# Effectiveness of an automated text message intervention for weight management in postpartum women with overweight or obesity (Supporting MumS (SMS)): a UK wide, multicentre, two arm, parallel group, randomised controlledti trial

**DOI:** 10.64898/2026.03.05.26347713

**Authors:** D Gallagher, E Spyreli, N Calder-MacPhee, K Crossley, C Feuillatre, A Ivory, B Karatas, CB Kelly, M Lind, E Osei-Asemani, R Potrick, H Stanton, S Bridges, E Coulman, C Free, P Hoddinott, AS Anderson, CR Cardwell, SU Dombrowski, S Heaney, F Kee, C McDowell, E McIntosh, L Murphy, JV Woodside, MC McKinley

## Abstract

**Objective:** To test the effectiveness of a postpartum behavioural intervention delivered by automated text messaging in reducing weight.

**Design:** Two parallel group, multicentre, randomised controlled trial.

**Setting:** Recruitment from five areas across the United Kingdom (Belfast, Bradford, Stirling, London and Cardiff) through healthcare and community pathways, including social media.

**Participants:** A diverse sample of 892 women between 6 weeks and 24 months postpartum, aged 18 years or more and with a body mass index of 25 kg/m^2^ or more, enrolled between May 2022 and May 2023: 445 were randomised to the intervention and 447 to an active control (comparator).

**Interventions:** 12 months of fully automated text messages with embedded behaviour change techniques and two-way messaging components to support weight loss and maintenance of weight loss in the postpartum period by targeting dietary, physical activity and weight management behaviours. The comparator group received 12 months of text messages on child health and development tailored to child age.

**Main outcome measures:** Primary outcome – weight in kilograms at 12 months (end of intervention). Secondary outcomes recorded at 6 and 12 months were changes in weight (at 6 months), body mass index, proportions of women with weight gain more than 5 kg, waist circumference, self-reported dietary intake, physical activity and infant feeding practices.

**Results:** 674 (75.6%) participants were included in the primary analysis. There was no difference found in the adjusted mean weight change between the intervention and active control groups (−0.1 kg (95% confidence interval -1.0 to 0.8, *P*= 0.84). Sensitivity analyses did not change these results. There was a small improvement in Fat and Fibre Barometer scores at 12 months in the intervention compared with control group (adjusted mean difference 0.09, 95% CI: 0.04 to 0.14; *P*<0.001) and an increase in physical activity scores (International Physical Activity Questionnaire – Short Form) at 12 months in the intervention group compared with the control group (adjusted mean difference 405.3 total MET minutes/week, 95% CI: 141.3 to 669.3; *P*= 0.003). At 12 months, infants in the intervention group consumed more eggs (adjusted mean difference 0.47 portions/week, 95% CI: 0.14 to 0.81; P= 0.01), beans/lentils (adjusted mean difference 0.40 portions/week, 95% CI: 0.09 to 0.71; P= 0.01), and nuts (adjusted mean difference 0.35 portions/week, 95% CI: 0.06 to 0.64; P= 0.02) than those in the control group.

**Conclusions:** A 12 month automated, interactive behavioural weight management intervention delivered by text message did not support weight loss for postpartum women but did have a positive impact on diet, physical activity and infant feeding behaviours.

**Trial registration:** ISRCTN Registry ISRCTN16299220

**WHAT IS ALREADY KNOWN ON THIS TOPIC:** Women desire support with self-care behaviours such as diet, physical activity and weight management in the postpartum period, but little support is currently available.

There is a lack of acceptable and effective weight management interventions designed for women during the postpartum period.

Most previous studies in the field have been affected by poor recruitment, high rates of attrition, lack of diversity and have not adequately considered the lived experience of women.

Mobile technologies can offer a more flexible and individualised ‘any time, any place’ approach to behavioural weight management interventions that may be well suited to this stage of life.

**WHAT THIS STUDY ADDS:** The trial methodology was highly acceptable to postpartum women, and recruitment and retention targets were met.

Overall, a fully automated two-way text message intervention did not support weight loss in a diverse sample of postpartum women from the United Kingdom but did support positive changes in diet, physical activity and infant feeding behaviours.

There was no differential effect of the intervention across socioeconomic or ethnic groups.

## Introduction

The health risks associated with obesity are well documented. Women have an increased risk of weight gain during the reproductive years (1). Maternal health data from England (2) indicated that in 2023/24 one in four women (26.2%) in early pregnancy had obesity with the highest prevalence noted in black women (36.3%) and those from the most deprived areas (32.4%). Furthermore, the prevalence of maternal obesity increased between first and subsequent pregnancies (2). Factors contributing to an upward weight trajectory at this stage of life are excess gestational weight gain, not losing gestational weight after pregnancy and further weight gain during the postpartum period (3–5), with heightened risk particularly between one and two years postpartum (6).

Postpartum weight management interventions could make a significant impact on reducing the long-term risks of obesity, heart disease, cancer and diabetes. Clinical guidance recommends that health professionals should advise women within two years of having a baby to eat a nutritious diet and keep physically active to encourage postpartum weight reduction (7, 8). However, women highlight a need for additional weight management support during the postpartum period as little is currently provided (9). The postpartum period is a physically, psychologically and emotionally demanding time for women (5). Factors such as physical recovery from pregnancy and childbirth, sleep deprivation, financial stress, mental health issues, caring responsibilities and a lack of time for self-care make this a uniquely challenging period for weight management (5). Careful consideration must, therefore, be given to the design of interventions for women at this stage of life (5).

Systematic review evidence indicates that behavioural interventions can effectively support postpartum weight management (10–12), particularly those that promote diet and physical activity behaviour change alongside self-monitoring (5). However, poor recruitment and retention and substantial heterogeneity and diversity in intervention designs makes it difficult to ascertain the most effective approaches (5, 10, 13, 14). To maximise engagement, the timing, setting and delivery format of postpartum interventions should seek to address specific barriers to health-related behaviour change that come with having a baby, such as childcare or time constraints (11, 15–18). What works at other stages of life may not necessarily be helpful in the postpartum period, as evidenced by the limited acceptability and success of in-person and structured weight management approaches that are incompatible with other demands on women’s time (19–21). Evidence, mostly from feasibility or pilot studies, indicates that technology usage holds promise as it can offer a more flexible approach to intervention delivery that may be more suited to the reality of women’s lives at this time (5, 22). For studies that have used technology to support postpartum women to achieve weight loss, it has often been used as an adjunct to in-person intervention delivery rather than the main modality, despite the barriers new mothers experience in engaging in-person (11); and small sample sizes and short term follow-up remain issues for these studies (22, 23).

Text message interventions can be cost-effective and readily scalable and have been used successfully to increase health-promoting behaviours including weight loss (24–29). Text messaging provides a simple communication mode that is accessible to most of the population supporting health equity (30–33). The Supporting MumS (SMS) trial was developed with consideration of the specific needs of women in the postpartum period in both the intervention and trial design (34, 35). The development, feasibility testing and cultural adaptation of the SMS intervention in preparation for a United Kingdom (UK) wide trial have been reported previously (35, 36). The 12-month SMS intervention adopts a fully automated approach to intervention delivery with two-way messaging to encourage engagement, delivery of specific behaviour change techniques (BCTs) and consideration of both weight loss and weight loss maintenance over the longer-term.

## Methods

### Trial design

A UK wide, multicentre, two arm, parallel group, superiority randomised controlled trial (RCT). Participant recruitment took place between April 2022 and May 2023 across five UK sites (Belfast, Northern Ireland; Stirling, Scotland; Cardiff, Wales; London, England; Bradford, England). Data collection at 12 months was completed by May 2024. The trial protocol, statistical analysis plan (SAP) and Process Evaluation Analysis Plan (PEAP) are provided in the Supplementary information and there were no major changes to the trial protocol after it commenced. The trial is reported here in accordance with the Consolidated Standards of Reporting Trials (CONSORT) 2025 checklist for RCTs (37). The study was prospectively registered on ISRCTN on 10/02/2022 (ISRCTN16299220; https://www.isrctn.com/ISRCTN16299220).

### Participants and recruitment

Eligible participants were women aged ≥18 years, who had given birth ≥6 weeks to ≤2 years prior, had a Body Mass Index (BMI) ≥25 kg/m^2^ and owned a mobile phone capable of receiving text messages (34). In line with National Institute for Health and Care Excellence (NICE) guideline NG1941(38), the term ’women’ was taken to include people who had been pregnant/given birth but did not identify as women. Exclusion criteria were: insufficient English language to understand short written messages; historical or planned weight loss surgery; diagnosis of anorexia nervosa or bulimia from a doctor; being on a specialist diet or receiving dietetic care; or participation in another weight management study or programme within the past three months. Pregnant women were not eligible, and participants who became pregnant during the trial discontinued, as the intervention was not designed for pregnancy.

Recruitment occurred across community and healthcare settings, reflecting potential real-world service access if implemented, as reported in detail elsewhere (39). Recruitment approaches were informed by the SMS pilot study (35), evidence on engaging minoritised ethnic and lower socioeconomic (SES) groups (21, 40), and using established researcher networks (39). Materials were developed with patient and public involvement (PPI) to ensure cultural relevance for different women (39).

### Randomisation and blinding

The allocation sequence was generated in Stata (using ralloc) by an independent statistician. Participants were randomised 1:1 to intervention or active control in blocks of four, stratified by site. The sequence was provided to the London School of Hygiene and Tropical Medicine (LSHTM) platform manager for upload to the secure web-based text messaging system. Following written informed consent and baseline data collection, participants were enrolled using the LSHTM online randomisation system linked to the platform. Participants became aware of allocation upon receiving text messages. Recruitment and outcome assessors were blinded, and participants were requested not to disclose their group at follow-up. Statistical analyses were conducted blinded until completion of the 12-month primary endpoint.

### Intervention and active control

Supporting MumS (SMS) intervention: intervention development and cultural adaptation with PPI, and the full intervention description according to the Template for Intervention Description and Replication guidance, are reported elsewhere (34–36). Briefly, the intervention is a fully automated, 12-month text message service, underpinned by the Health Action Process Approach (HAPA) (41) and behaviour maintenance theories (42). It consists of a library of text messages focused on diet and physical activity behaviours with embedded BCTs known to be positively associated with weight loss and weight loss maintenance. Developed with PPI for postpartum women, it is self-directed, offering a range of evidence-based health-promoting strategies, allowing women to choose what works for them. Messages adopt a friendly, non-pressurising tone, signpost to external credible resources, provide encouragement and motivation, discourage guilt and promote self-reflection (34, 35). A core library of 353 text messages delivered over 12 months (14–15/week in months one to two, 9–10/week in months three to six, 4–5/week in months 7-12) includes: weekly bidirectional prompts to self-weigh and report weight (n=50); “Yes/No” questions (n=36) encouraging engagement, self-monitoring, relapse prevention and providing feedback, with automated tailored replies; and on-demand ‘trigger words’ (‘slip-up’; ‘crave; ‘bad day’ or ‘tired’) that when texted generate automated tailored replies addressing barrier management and relapse prevention. A sample of messages delivered over a two week period (weeks 18 and 19 of the intervention) is presented in Supplementary Table 1. From months 7-12, when the intervention focus shifts from initiation to maintenance, weekly weight prompts asked participants to report weight alongside ‘up’ or ‘down’ or ‘same’ comparing their weight with the previous week, triggering automated, tailored feedback. All automated responses are drawn from a bank of pre-programmed messages.

Active control: Participants receive three weekly unidirectional text messages on child health and development for 12 months, aligned with their baby’s age (entered at baseline). Messages exclude intervention content (i.e. diet, physical activity, weight management, and BCTs). After 12-month follow-up, active control participants are offered a booklet summarising the intervention.

### Data collection

Data were collected at baseline (post-consent, pre-randomisation), and at 6 and 12 months post-randomisation (end of intervention). Trained researchers measured height, weight and waist circumference using standardised protocols and calibrated scales. Researchers recorded baseline demographics in the Case Report Form (CRF). Participants completed self-report questionnaires on paper (given at visit with paid return envelope) or online (Qualtrics survey platform) according to preference at each visit. Researchers assisted with paper questionnaires during visits or via telephone if needed. To maximise data completeness, researchers followed up on non-returned questionnaires, and all data were checked, and queries (e.g., missing data, invalid ranges etc) and actions to resolve them were logged, with participant clarification where required.

Trial methods prioritised participant retention through flexible data collection and engagement strategies. To facilitate women’s participation, assessments could be conducted via home visits or at a participant-chosen venue e.g. university/community setting. Preferred contact methods were documented and prioritised, and visits were scheduled at convenient times for the participant with reminders sent one month, one week and one day in advance. Up to three contact attempts were made to arrange visits. If a face-to-face 12-month visit (primary endpoint) had not been arranged after contact attempts were exhausted, participants were asked to self-report weight and complete an online questionnaire. Participants received a £25 voucher on receipt of completed questionnaires at each timepoint (0, 6 and 12 months), as a token of appreciation for their time committed to trial assessments. Those lost to follow-up at one timepoint but not withdrawn were invited to attend future visits.

### Outcomes

Primary and secondary outcomes were: Primary - mean weight change (kg) from baseline to 12 months; Secondary – mean weight change (kg) from baseline to 6 months, and mean waist circumference (cm) change from baseline, proportions of women gaining a substantial amount of weight (>5kg) from baseline, change in dietary intake (Fat and Fibre Barometer (FFB) (43)), frequency of consuming common sugary foods and alcohol), change in physical activity including sedentary behaviour (sitting) (International Physical Activity Questionnaire-Short Form (IPAQ-SF) (44)) and infant feeding practices (based on the UK Infant Feeding Survey 2010 (45)), at 6 and 12 months, respectively.

### Acceptability of trial methods

Assessment of acceptability of the trials methods for participants is summarised in the Supplementary information.

### Fidelity of researcher blinding

To assess whether blinding was maintained, any instances where researchers became aware of participants’ group allocation or message receipt were reported to the trial manager and recorded.

### Adverse events

Serious adverse events (SAEs) were recorded from consent until one month post-intervention (12-months). Site leads reviewed and classified events by severity, causality and expectedness, following Good Clinical Practice guidelines and Sponsor procedures.

### Sample size

The target sample size was 888 women over 12 months, with site-specific targets (London and Bradford: n=189 each; Belfast, Stirling and Cardiff: n=170 each). Pregnancy-related withdrawals were not replaced unless thresholds for loss to follow-up (12%) or pregnancy rates (15%) observed in the pilot RCT (35) were exceeded at 6 months, as monitored by the Project Management Team.

In the SMS pilot trial (35), the intervention group lost a mean 1.75 kg weight versus a 0.19 kg gain in the active control group between baseline and 12 months, yielding an adjusted between-group difference of -1.67 kg (95% CI -4.88 to 1.55). Using these data and a standard deviation of 7.5 kg, 594 completing participants (297 per group) provide >90% power to detect a 2 kg difference between groups in mean weight change from baseline, at the 5% significance level. A 2 kg threshold is associated with metabolic benefits and often used to power weight loss studies (46).

Allowing for a 15% pregnancy dropout rate (35) and evidence of higher rates in more diverse populations (∼22% in the Born in Bradford’s Better Start (BiBBS) cohort (47)), sample sizes were inflated to achieve 119 completing participants per site. This required 170 participants in Belfast, Stirling and Cardiff (15% pregnancy rate + 15% loss to follow-up) and 189 in Bradford and London (22% pregnancy rate + 15% loss to follow-up), giving a total target sample of 888 women (444 per group).

### Statistical analysis

A SAP was made available prior to commencing analysis (https://doi.org/10.1186/ISRCTN16299220) (see Supplementary information). A CONSORT flow diagram (37) illustrates participant progress through allocation, follow-up and analysis. Trial enrolment has been reported elsewhere (39). Retention by group and timepoint was examined, including numbers/proportions excluded due to pregnancy or discontinuation of text messages. Baseline characteristics of the randomised and primary analysis samples were compared by group using descriptive statistics and frequencies.

Weight change from baseline to each follow-up was calculated, and women were categorised as gaining >5kg at 6 and 12 months. Validated scales were scored per recommendations: FFB mean scores (43), and IPAQ-SF MET minutes/week, further categorised into activity levels (low/moderate/high), alongside minutes sitting per weekday (44)). Mean frequency of sugary food intake (across four food types) was calculated. Infant feeding data were collected on frequency (‘more than once a day’ to ‘never’) of giving 21 food types (45) and converted to estimated weekly portions (e.g. ‘once a day’ converted to ‘7 portions/week).

Primary, secondary and exploratory outcomes were analysed using an intention to treat approach, including all randomised participants with available endpoint data, comparing intervention and active control groups. Tests were two-sided at the 5% significance level, with mean weight change (kg) at 12 months considered the primary analysis. All subsequent tests were considered descriptive/exploratory, with no correction for multiple testing.

Analysis of covariance (ANCOVA) was used to analyse the primary outcome and continuous secondary outcomes, adjusting for baseline values, site, ethnicity, and recruitment pathway (healthcare vs community). Model assumptions were assessed using residual plots and histograms. Baseline and endpoint group means are presented alongside adjusted mean differences with 95% confidence intervals (CIs) and p-values. Secondary categorical outcomes were prespecified as binary; however, some categorical outcomes (IPAQ-SF physical activity levels and alcohol consumption) were analysed as ordinal variables where more appropriate, using the same approach as for binary outcomes. Binary and ordinal outcomes were analysed using logistic regression and proportional odds models, respectively, adjusting for site, recruitment pathway and ethnicity. The proportional odds assumption was assessed using the test of parallel lines. Adjusted odds ratios, 95% CIs and corresponding *P*-values are reported.

Prespecified sensitivity analyses (see the SAP in Supplementary information) repeated the primary analysis by including self-reported 12-month weights and accounting for missing data using: 1) baseline observation carried forward; 2) last observation carried forward; and 3) multiple imputation. Ten imputed datasets were created, with models including treatment site, recruitment pathway, ethnicity and baseline weight. Aδ-based sensitivity analysis (δ= 0.9 kg) (48) assumed individuals with missing data were, on average, 0.9 kg heavier than expected; ten imputed datasets were again created with the same covariates. An exploratory post hoc analysis excluded participants reporting taking medications or undergoing surgery for weight loss during the trial.

Prespecified exploratory subgroup analyses examined differential intervention effects on the primary outcome, and secondary diet (FFB mean scores (43)) and physical activity (IPAQ MET minutes/week (44)) outcomes by site, recruitment pathway (NHS or community), ethnicity, index of multiple deprivation (IMD) quintile, employment, highest education, income (<£30,000 or ≥£30,001 or don’t know), BMI at study entry, parity, and weeks postpartum at study entry. Interaction tests were conducted separately by including interaction terms within ANCOVA regression models. Differential intervention effects on 12-month weight across sites were presented in a forest plot.

### Trial methods analysis

The analysis of trial methods acceptability is outlined in the Supplementary information.

Numbers of instances of researcher unblinding, along with details of the unblinding, were summarised.

Numbers of SAEs were summarised, and the categorisation and onward reporting of SAEs detailed.

All quantitative analyses were performed using Stata version 16 (Statcorp, USA) and IBM SPSS version 29 software.

#### Patient and public involvement

PPI was essential and integral across all stages of this trial, with representatives and women with lived experience involved throughout the research cycle. Intervention development, cultural adaptation and trial design, with embedded PPI, are reported elsewhere (34–36) including designing the intervention and trial approaches with cognisance of addressing participation barriers and minimising burden. PPI contributors sit on the Project Management Team and Trial Steering Committee, informing decision making, and are involved in reviewing dissemination materials, including co-authoring publications. Participant preferences for receiving results, captured at 24 months (to be reported elsewhere), will guide dissemination to non-academic audiences.

## Results

### Participant characteristics and retention

Between April 2022 and May 2023, 2457 women expressed an interest in the study, 1227 were screened and 892 were enrolled and randomised (intervention 445; active control 447) (Figure 1). Trial recruitment and baseline characteristics of the sample were reported previously (39). Women were from all four UK countries, 301 (33.6%) were from minoritised ethnic groups and 477 (53.5%) lived in the two most deprived IMD quintile areas. Most had obesity (547, 61.3%)) versus overweight (345, 38.7%). At study entry, 414 (46.4%) had an Edinburgh Postnatal Depression Scale (EPDS) score of 9 or above and 245 (27.4%) had a long-term physical or mental health condition. Participants spanned the extended postpartum inclusion period (≤6 months= 298, 33.4%; 7-12 months postpartum= 254, 28.5%; >12 months postpartum= 340, 38.1%). English was a second language for 104 women (intervention 45; active control 59), and 15 had a recent multiple pregnancy (twins 14; triplets 1).

**Figure 1:**
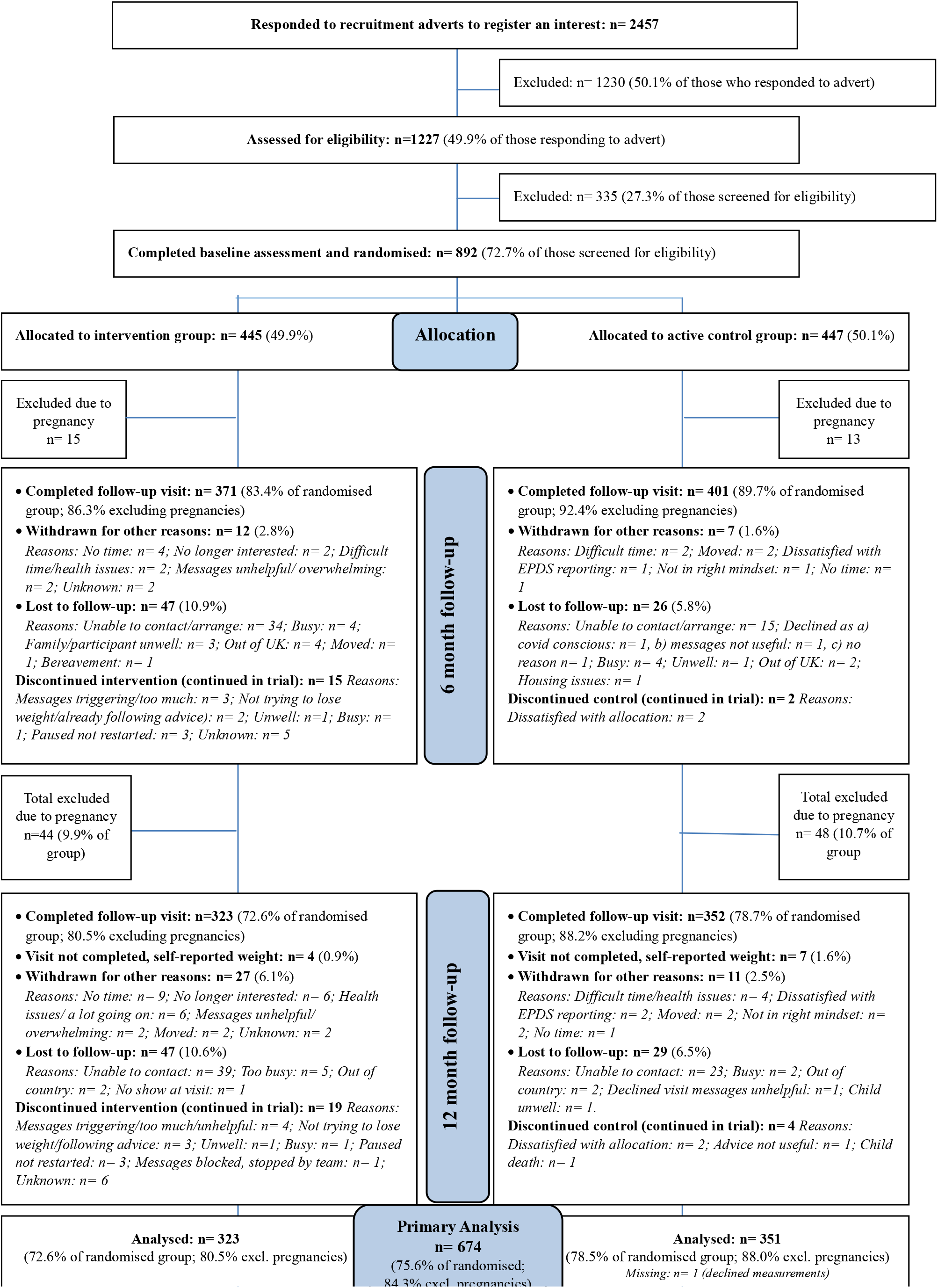
CONSORT flowchart.

Data collection visits were completed for 772 (86.5%) and 675 (75.7%) participants at 6 months (November 2022-October 2023) and 12 months (May 2023-April 2024), respectively (Figure 1). Excluding pregnancies (women stopped participation if pregnant), retention was 89.4% at 6 months and 84.4% at 12 months. Pregnancy exclusion was 3% at 6 months and 10% by 12 months. Target sample sizes at follow-up were exceeded and were 6% higher for the control group (6 months, 83.4% vs 89.7%; 12 months, 72.6% vs 78.7%). At 12 months, one participant declined weight measurement and was excluded from the primary analysis, and 11 (1.2%) provided self-reported weight (Figure 1).

Retention did not notably differ by participant characteristics; baseline characteristics were comparable between the randomised groups and those included in the primary analysis (Table 1).

**Table 1:**
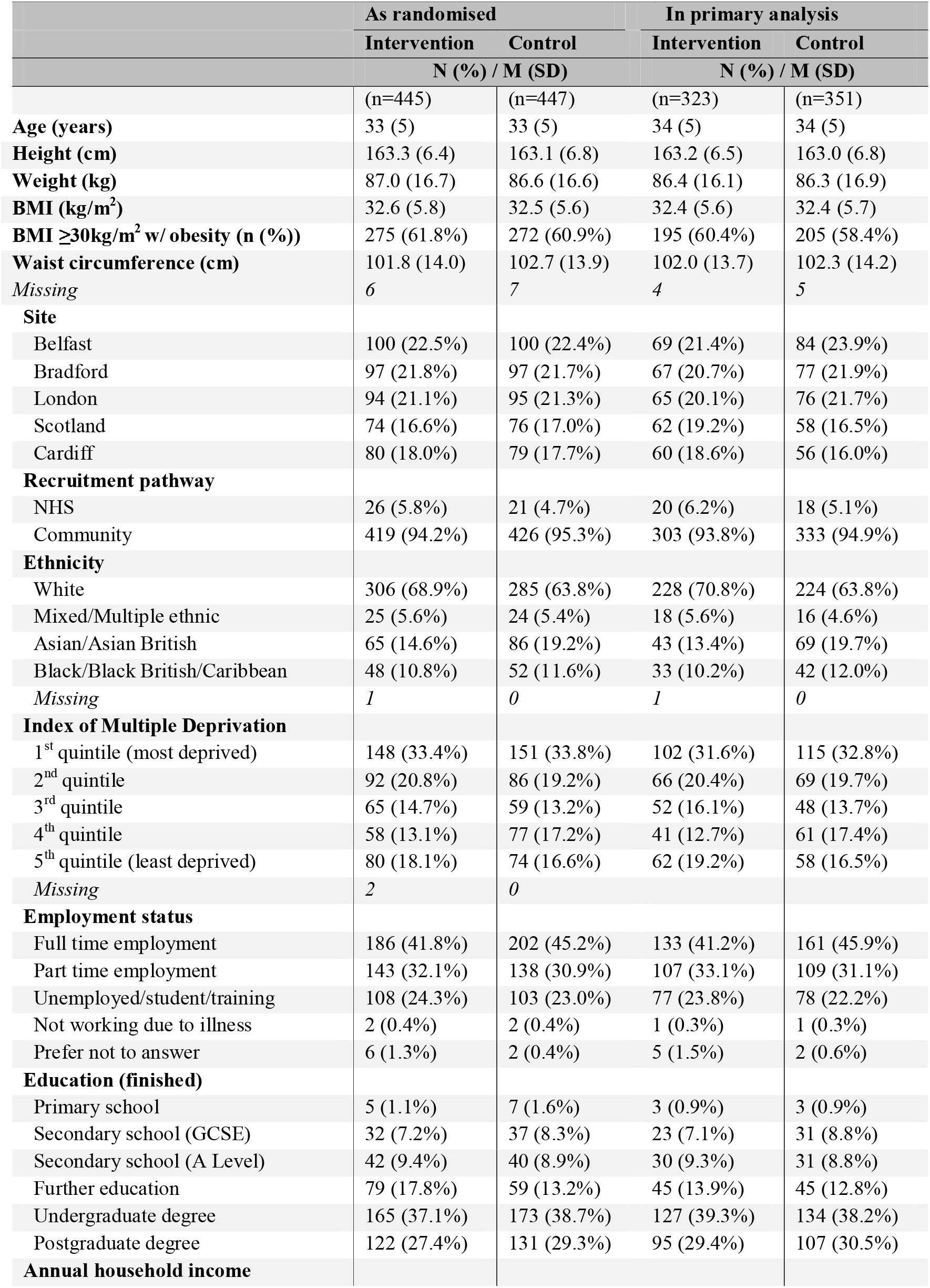
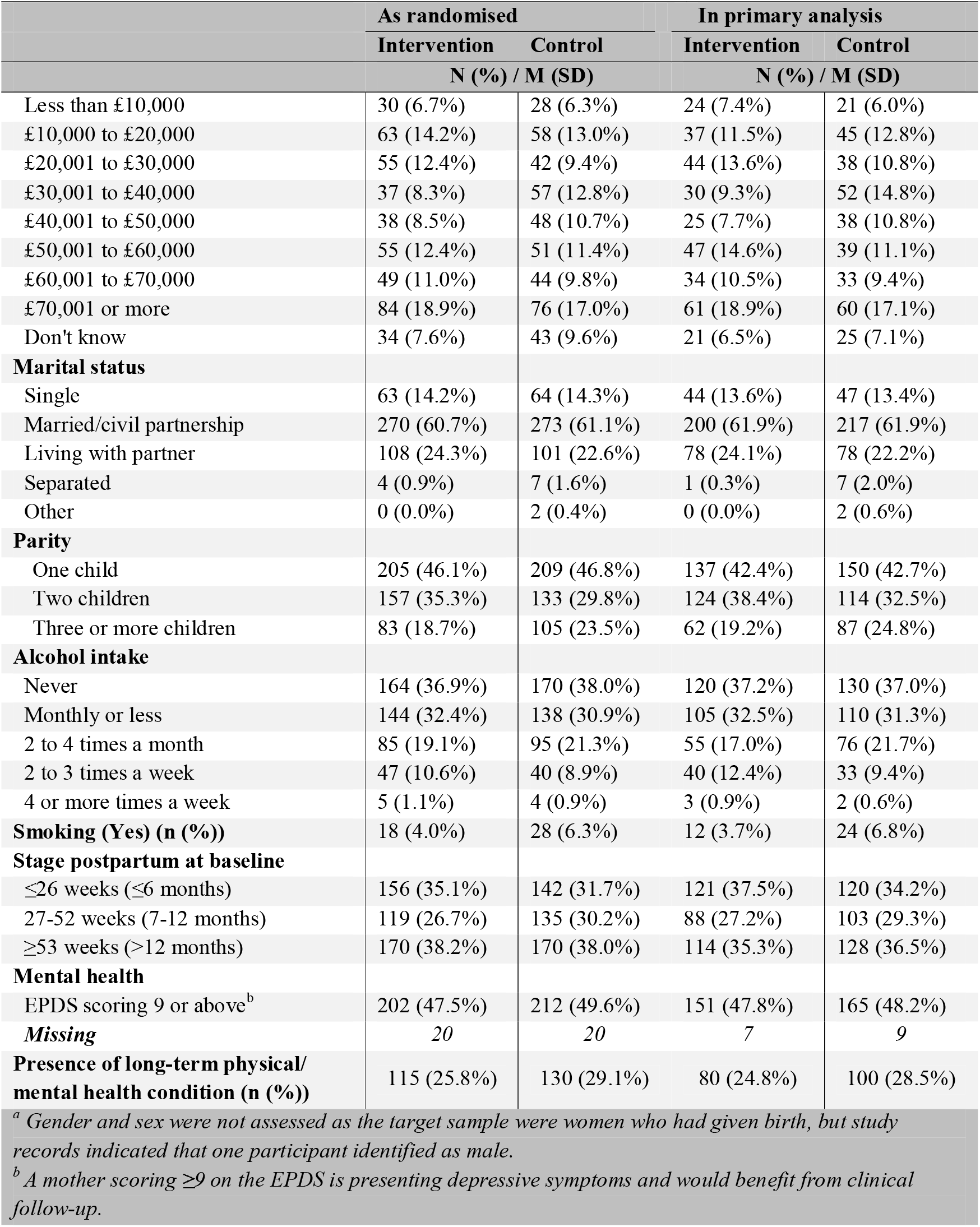
Baseline characteristics comparing randomised and primary analysis samples by group.

|  | As randomised |  | In primary analysis |  |
| --- | --- | --- | --- | --- |
|  | Intervention | Control | Intervention | Control |
|  | N (%) / M (SD) |  | N (%) / M (SD) |  |
|  | (n=445) | (n=447) | (n=323) | (n=351) |
| **Age (years)** | 33 (5) | 33 (5) | 34 (5) | 34 (5) |
| **Height (cm)** | 163.3 (6.4) | 163.1 (6.8) | 163.2 (6.5) | 163.0 (6.8) |
| **Weight (kg)** | 87.0 (16.7) | 86.6 (16.6) | 86.4 (16.1) | 86.3 (16.9) |
| **BMI (kg/m<sup>2</sup>)** | 32.6 (5.8) | 32.5 (5.6) | 32.4 (5.6) | 32.4 (5.7) |
| **BMI $\geq 30$ kg/m<sup>2</sup> w/ obesity (n (%))** | 275 (61.8%) | 272 (60.9%) | 195 (60.4%) | 205 (58.4%) |
| **Waist circumference (cm)** | 101.8 (14.0) | 102.7 (13.9) | 102.0 (13.7) | 102.3 (14.2) |
| *Missing* | 6 | 7 | 4 | 5 |
| **Site** |  |  |  |  |
| Belfast | 100 (22.5%) | 100 (22.4%) | 69 (21.4%) | 84 (23.9%) |
| Bradford | 97 (21.8%) | 97 (21.7%) | 67 (20.7%) | 77 (21.9%) |
| London | 94 (21.1%) | 95 (21.3%) | 65 (20.1%) | 76 (21.7%) |
| Scotland | 74 (16.6%) | 76 (17.0%) | 62 (19.2%) | 58 (16.5%) |
| Cardiff | 80 (18.0%) | 79 (17.7%) | 60 (18.6%) | 56 (16.0%) |
| **Recruitment pathway** |  |  |  |  |
| NHS | 26 (5.8%) | 21 (4.7%) | 20 (6.2%) | 18 (5.1%) |
| Community | 419 (94.2%) | 426 (95.3%) | 303 (93.8%) | 333 (94.9%) |
| **Ethnicity** |  |  |  |  |
| White | 306 (68.9%) | 285 (63.8%) | 228 (70.8%) | 224 (63.8%) |
| Mixed/Multiple ethnic | 25 (5.6%) | 24 (5.4%) | 18 (5.6%) | 16 (4.6%) |
| Asian/Asian British | 65 (14.6%) | 86 (19.2%) | 43 (13.4%) | 69 (19.7%) |
| Black/Black British/Caribbean | 48 (10.8%) | 52 (11.6%) | 33 (10.2%) | 42 (12.0%) |
| *Missing* | 1 | 0 | 1 | 0 |
| **Index of Multiple Deprivation** |  |  |  |  |
| 1<sup>st</sup> quintile (most deprived) | 148 (33.4%) | 151 (33.8%) | 102 (31.6%) | 115 (32.8%) |
| 2<sup>nd</sup> quintile | 92 (20.8%) | 86 (19.2%) | 66 (20.4%) | 69 (19.7%) |
| 3<sup>rd</sup> quintile | 65 (14.7%) | 59 (13.2%) | 52 (16.1%) | 48 (13.7%) |
| 4<sup>th</sup> quintile | 58 (13.1%) | 77 (17.2%) | 41 (12.7%) | 61 (17.4%) |
| 5<sup>th</sup> quintile (least deprived) | 80 (18.1%) | 74 (16.6%) | 62 (19.2%) | 58 (16.5%) |
| *Missing* | 2 | 0 |  |  |
| **Employment status** |  |  |  |  |
| Full time employment | 186 (41.8%) | 202 (45.2%) | 133 (41.2%) | 161 (45.9%) |
| Part time employment | 143 (32.1%) | 138 (30.9%) | 107 (33.1%) | 109 (31.1%) |
| Unemployed/student/training | 108 (24.3%) | 103 (23.0%) | 77 (23.8%) | 78 (22.2%) |
| Not working due to illness | 2 (0.4%) | 2 (0.4%) | 1 (0.3%) | 1 (0.3%) |
| Prefer not to answer | 6 (1.3%) | 2 (0.4%) | 5 (1.5%) | 2 (0.6%) |
| **Education (finished)** |  |  |  |  |
| Primary school | 5 (1.1%) | 7 (1.6%) | 3 (0.9%) | 3 (0.9%) |
| Secondary school (GCSE) | 32 (7.2%) | 37 (8.3%) | 23 (7.1%) | 31 (8.8%) |
| Secondary school (A Level) | 42 (9.4%) | 40 (8.9%) | 30 (9.3%) | 31 (8.8%) |
| Further education | 79 (17.8%) | 59 (13.2%) | 45 (13.9%) | 45 (12.8%) |
| Undergraduate degree | 165 (37.1%) | 173 (38.7%) | 127 (39.3%) | 134 (38.2%) |
| Postgraduate degree | 122 (27.4%) | 131 (29.3%) | 95 (29.4%) | 107 (30.5%) |
| **Annual household income** |  |  |  |  |
|  | Intervention | Control | Intervention | Control |
|  | N (%) / M (SD) |  | N (%) / M (SD) |  |
| Less than £10,000 | 30 (6.7%) | 28 (6.3%) | 24 (7.4%) | 21 (6.0%) |
| £10,000 to £20,000 | 63 (14.2%) | 58 (13.0%) | 37 (11.5%) | 45 (12.8%) |
| £20,001 to £30,000 | 55 (12.4%) | 42 (9.4%) | 44 (13.6%) | 38 (10.8%) |
| £30,001 to £40,000 | 37 (8.3%) | 57 (12.8%) | 30 (9.3%) | 52 (14.8%) |
| £40,001 to £50,000 | 38 (8.5%) | 48 (10.7%) | 25 (7.7%) | 38 (10.8%) |
| £50,001 to £60,000 | 55 (12.4%) | 51 (11.4%) | 47 (14.6%) | 39 (11.1%) |
| £60,001 to £70,000 | 49 (11.0%) | 44 (9.8%) | 34 (10.5%) | 33 (9.4%) |
| £70,001 or more | 84 (18.9%) | 76 (17.0%) | 61 (18.9%) | 60 (17.1%) |
| Don't know | 34 (7.6%) | 43 (9.6%) | 21 (6.5%) | 25 (7.1%) |
| **Marital status** |  |  |  |  |
| Single | 63 (14.2%) | 64 (14.3%) | 44 (13.6%) | 47 (13.4%) |
| Married/civil partnership | 270 (60.7%) | 273 (61.1%) | 200 (61.9%) | 217 (61.9%) |
| Living with partner | 108 (24.3%) | 101 (22.6%) | 78 (24.1%) | 78 (22.2%) |
| Separated | 4 (0.9%) | 7 (1.6%) | 1 (0.3%) | 7 (2.0%) |
| Other | 0 (0.0%) | 2 (0.4%) | 0 (0.0%) | 2 (0.6%) |
| **Parity** |  |  |  |  |
| One child | 205 (46.1%) | 209 (46.8%) | 137 (42.4%) | 150 (42.7%) |
| Two children | 157 (35.3%) | 133 (29.8%) | 124 (38.4%) | 114 (32.5%) |
| Three or more children | 83 (18.7%) | 105 (23.5%) | 62 (19.2%) | 87 (24.8%) |
| **Alcohol intake** |  |  |  |  |
| Never | 164 (36.9%) | 170 (38.0%) | 120 (37.2%) | 130 (37.0%) |
| Monthly or less | 144 (32.4%) | 138 (30.9%) | 105 (32.5%) | 110 (31.3%) |
| 2 to 4 times a month | 85 (19.1%) | 95 (21.3%) | 55 (17.0%) | 76 (21.7%) |
| 2 to 3 times a week | 47 (10.6%) | 40 (8.9%) | 40 (12.4%) | 33 (9.4%) |
| 4 or more times a week | 5 (1.1%) | 4 (0.9%) | 3 (0.9%) | 2 (0.6%) |
| **Smoking (Yes) (n (%))** | 18 (4.0%) | 28 (6.3%) | 12 (3.7%) | 24 (6.8%) |
| **Stage postpartum at baseline** |  |  |  |  |
| ≤26 weeks (≤6 months) | 156 (35.1%) | 142 (31.7%) | 121 (37.5%) | 120 (34.2%) |
| 27-52 weeks (7-12 months) | 119 (26.7%) | 135 (30.2%) | 88 (27.2%) | 103 (29.3%) |
| ≥53 weeks (>12 months) | 170 (38.2%) | 170 (38.0%) | 114 (35.3%) | 128 (36.5%) |
| **Mental health** |  |  |  |  |
| EPDS scoring 9 or above<sup>b</sup> | 202 (47.5%) | 212 (49.6%) | 151 (47.8%) | 165 (48.2%) |
| **Missing** | 20 | 20 | 7 | 9 |
| **Presence of long-term physical/mental health condition (n (%))** | 115 (25.8%) | 130 (29.1%) | 80 (24.8%) | 100 (28.5%) |

<sup>a</sup> Gender and sex were not assessed as the target sample were women who had given birth, but study records indicated that one participant identified as male.
<sup>b</sup> A mother scoring ≥9 on the EPDS is presenting depressive symptoms and would benefit from clinical follow-up.

### Main findings

The primary outcome of weight in kg at 12 months was similar between the intervention and control groups (adjusted mean difference -0.1 kg, 95% CI: -1.0 to 0.8, *P*= 0.84) (Table 2). For secondary anthropometric outcomes at 12 months, there were no differences between groups found for BMI (adjusted mean difference -0.1 kg/m^2^, 95% CI: -0.4 to 0.3, *P*= 0.73), waist circumference (adjusted mean difference -0.5 cm, 95% CI: -1.5 to 0.6, *P*= 0.40) or the proportions of women gaining >5 kg (12% vs 11%; adjusted OR 0.86, 95% CI: 0.53 to 1.40, *P*= 0.55). Results were similar at 6 months and remained unchanged in the sensitivity analysis (Table 2).

For secondary diet and activity outcomes, FFB scores were improved for the intervention group compared with control group (adjusted mean difference 0.09, 95% CI: 0.04 to 0.14; *P* <0.001) but no between group differences found for intake of sugary foods or alcohol (Table 3). Physical activity (IPAQ MET minutes/week) increased for the intervention group compared with the control group (adjusted mean difference 405.3 MET minutes/week, 95% CI: 141.3 to 669.3; *P*= 0.003) with a notable shift from low to moderate or high activity levels in the intervention group relative to control (adjusted OR 1.41, 95% CI: 1.05 to 1.90; *P*= 0.02), but no differences found for sitting time (Table 3). Similar trends for diet and activity were observed at 6 months, with an additional reduction in intake of sugary foods found for the intervention group compared with control (adjusted mean difference -0.11, 95% CI: -0.20 to -0.01; *P*= 0.04) (Table 3).

For infant feeding items (Supplementary Tables 2 and 3), increased weekly portions of eggs, beans/lentils and nuts were given to infants in the intervention group compared with the control group at 12 months (eggs-adjusted mean difference 0.47 portions/week, 95% CI: 0.14 to 0.81; *P=* 0.01; beans/lentils-adjusted mean difference 0.40 portions/week, 95% CI: 0.09 to 0.71; *P=* 0.01; nuts-adjusted mean difference 0.35 portions/week, 95% CI: 0.06 to 0.64; *P=* 0.02).

Prespecified sub-group analyses for primary (weight) (Table 4) and secondary outcomes (diet: FFB scores; physical activity: IPAQ MET minutes per week) showed consistent results across sub-groups including IMD, ethnicity and site (Supplementary Tables 4 and 5). A forest plot showing the adjusted mean difference in weight (and 95% CIs) between groups at 12 months by site is shown in Supplementary Figure 1.

### Primary and secondary anthropometric measures

**Table 2:**
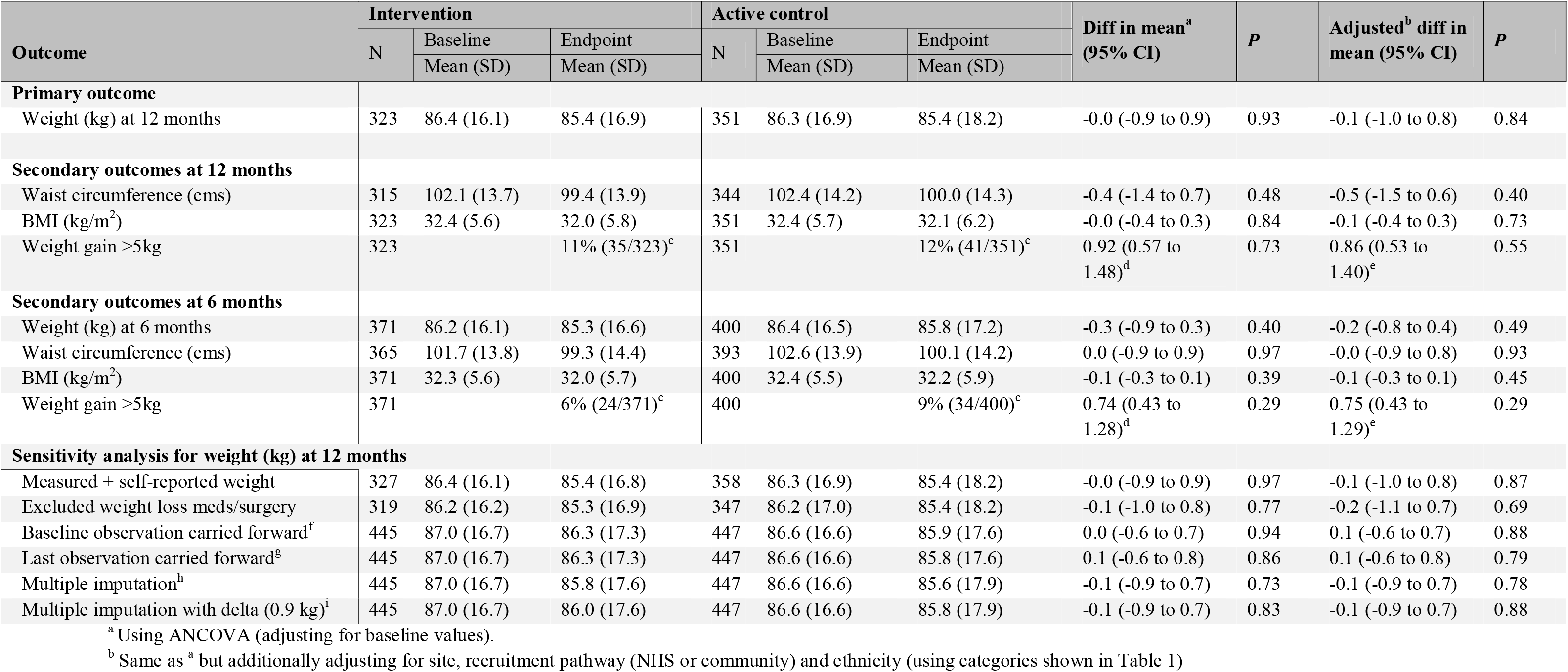
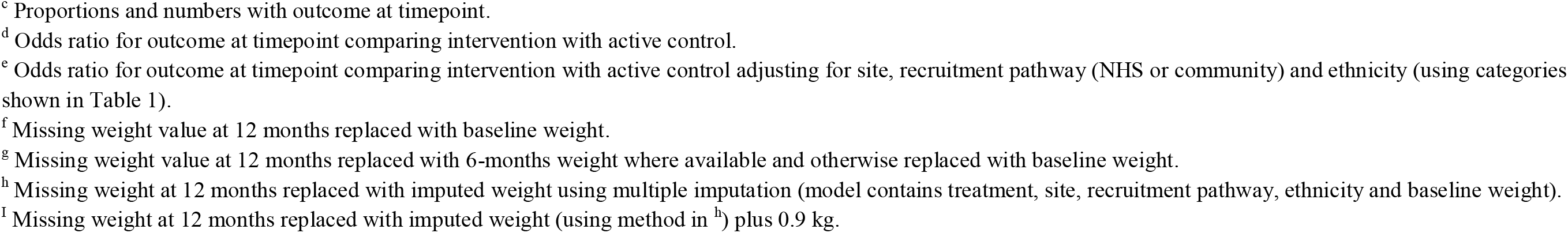
Comparisons of weight and body composition outcomes between intervention and active control at 6 and 12 months, including sensitivity analysis.

| Outcome | Intervention |  |  | Active control |  |  | Diff in mean<sup>a</sup><br>(95% CI) | P | Adjusted<sup>b</sup> diff in<br>mean (95% CI) | P |
| --- | --- | --- | --- | --- | --- | --- | --- | --- | --- | --- |
|  | N | Baseline | Endpoint | N | Baseline | Endpoint |  |  |  |  |
|  |  | Mean (SD) | Mean (SD) |  | Mean (SD) | Mean (SD) |  |  |  |  |
| **Primary outcome** |  |  |  |  |  |  |  |  |  |  |
| Weight (kg) at 12 months | 323 | 86.4 (16.1) | 85.4 (16.9) | 351 | 86.3 (16.9) | 85.4 (18.2) | -0.0 (-0.9 to 0.9) | 0.93 | -0.1 (-1.0 to 0.8) | 0.84 |
| **Secondary outcomes at 12 months** |  |  |  |  |  |  |  |  |  |  |
| Waist circumference (cms) | 315 | 102.1 (13.7) | 99.4 (13.9) | 344 | 102.4 (14.2) | 100.0 (14.3) | -0.4 (-1.4 to 0.7) | 0.48 | -0.5 (-1.5 to 0.6) | 0.40 |
| BMI (kg/m<sup>2</sup>) | 323 | 32.4 (5.6) | 32.0 (5.8) | 351 | 32.4 (5.7) | 32.1 (6.2) | -0.0 (-0.4 to 0.3) | 0.84 | -0.1 (-0.4 to 0.3) | 0.73 |
| Weight gain >5kg | 323 |  | 11% (35/323)<sup>c</sup> | 351 |  | 12% (41/351)<sup>c</sup> | 0.92 (0.57 to 1.48)<sup>d</sup> | 0.73 | 0.86 (0.53 to 1.40)<sup>e</sup> | 0.55 |
| **Secondary outcomes at 6 months** |  |  |  |  |  |  |  |  |  |  |
| Weight (kg) at 6 months | 371 | 86.2 (16.1) | 85.3 (16.6) | 400 | 86.4 (16.5) | 85.8 (17.2) | -0.3 (-0.9 to 0.3) | 0.40 | -0.2 (-0.8 to 0.4) | 0.49 |
| Waist circumference (cms) | 365 | 101.7 (13.8) | 99.3 (14.4) | 393 | 102.6 (13.9) | 100.1 (14.2) | 0.0 (-0.9 to 0.9) | 0.97 | -0.0 (-0.9 to 0.8) | 0.93 |
| BMI (kg/m<sup>2</sup>) | 371 | 32.3 (5.6) | 32.0 (5.7) | 400 | 32.4 (5.5) | 32.2 (5.9) | -0.1 (-0.3 to 0.1) | 0.39 | -0.1 (-0.3 to 0.1) | 0.45 |
| Weight gain >5kg | 371 |  | 6% (24/371)<sup>c</sup> | 400 |  | 9% (34/400)<sup>c</sup> | 0.74 (0.43 to 1.28)<sup>d</sup> | 0.29 | 0.75 (0.43 to 1.29)<sup>e</sup> | 0.29 |
| **Sensitivity analysis for weight (kg) at 12 months** |  |  |  |  |  |  |  |  |  |  |
| Measured + self-reported weight | 327 | 86.4 (16.1) | 85.4 (16.8) | 358 | 86.3 (16.9) | 85.4 (18.2) | -0.0 (-0.9 to 0.9) | 0.97 | -0.1 (-1.0 to 0.8) | 0.87 |
| Excluded weight loss meds/surgery | 319 | 86.2 (16.2) | 85.3 (16.9) | 347 | 86.2 (17.0) | 85.4 (18.2) | -0.1 (-1.0 to 0.8) | 0.77 | -0.2 (-1.1 to 0.7) | 0.69 |
| Baseline observation carried forward<sup>f</sup> | 445 | 87.0 (16.7) | 86.3 (17.3) | 447 | 86.6 (16.6) | 85.9 (17.6) | 0.0 (-0.6 to 0.7) | 0.94 | 0.1 (-0.6 to 0.7) | 0.88 |
| Last observation carried forward<sup>g</sup> | 445 | 87.0 (16.7) | 86.3 (17.3) | 447 | 86.6 (16.6) | 85.8 (17.6) | 0.1 (-0.6 to 0.8) | 0.86 | 0.1 (-0.6 to 0.8) | 0.79 |
| Multiple imputation<sup>h</sup> | 445 | 87.0 (16.7) | 85.8 (17.6) | 447 | 86.6 (16.6) | 85.6 (17.9) | -0.1 (-0.9 to 0.7) | 0.73 | -0.1 (-0.9 to 0.7) | 0.78 |
| Multiple imputation with delta (0.9 kg)<sup>i</sup> | 445 | 87.0 (16.7) | 86.0 (17.6) | 447 | 86.6 (16.6) | 85.8 (17.9) | -0.1 (-0.9 to 0.7) | 0.83 | -0.1 (-0.9 to 0.7) | 0.88 |

<sup>a</sup> Using ANCOVA (adjusting for baseline values).
<sup>b</sup> Same as <sup>a</sup> but additionally adjusting for site, recruitment pathway (NHS or community) and ethnicity (using categories shown in Table 1)
<sup>c</sup> Proportions and numbers with outcome at timepoint.
<sup>d</sup> Odds ratio for outcome at timepoint comparing intervention with active control.
<sup>e</sup> Odds ratio for outcome at timepoint comparing intervention with active control adjusting for site, recruitment pathway (NHS or community) and ethnicity (using categories shown in Table 1).
<sup>f</sup> Missing weight value at 12 months replaced with baseline weight.
<sup>g</sup> Missing weight value at 12 months replaced with 6-months weight where available and otherwise replaced with baseline weight.
<sup>h</sup> Missing weight at 12 months replaced with imputed weight using multiple imputation (model contains treatment, site, recruitment pathway, ethnicity and baseline weight).
<sup>i</sup> Missing weight at 12 months replaced with imputed weight (using method in <sup>h</sup>) plus 0.9 kg.

### Secondary outcomes – diet (including alcohol) and physical activity (including sedentary behaviour)

**Table 3:** Comparisons of secondary outcomes between intervention and active control at 6 and 12 months.

|  | Intervention |  |  | Active Control |  |  | Diff in mean<sup>a</sup><br>(95% CI) | P | Adjusted<sup>b</sup> diff in<br>mean (95 % CI) | P |
| --- | --- | --- | --- | --- | --- | --- | --- | --- | --- | --- |
|  | n | Baseline<br>Mean (SD) | Endpoint<br>Mean (SD) | N | Baseline<br>Mean (SD) | Endpoint<br>Mean (SD) |  |  |  |  |
| **Outcomes at 12 months** |  |  |  |  |  |  |  |  |  |  |
| Diet (FFB scores) | 303 | 3.03 (0.4) | 3.20 (0.4) | 335 | 3.09 (0.4) | 3.15 (0.4) | **0.09 (0.04 to 0.14)** | **<0.001** | **0.09 (0.04 to 0.14)** | **<0.001** |
| Diet (sugary foods intake) | 303 | 2.24 (0.9) | 1.91 (0.8) | 332 | 2.18 (0.9) | 1.92 (0.8) | -0.05 (-0.15 to 0.05) | 0.33 | -0.06 (-0.17 to 0.04) | 0.22 |
| Physical Activity (IPAQ-SF MET<br>minutes/week) | 297 | 1430.0<br>(1372.7) | 2033.5<br>(1928.7) | 331 | 1612.0<br>(1769.2) | 1712.4<br>(1781.6) | **414.8**<br>**(153.3 to 676.2)** | **0.002** | **405.3**<br>**(141.3 to 669.3)** | **0.003** |
| Sitting (IPAQ-SF minutes/<br>weekday) | 221 | 332.3 (172.3) | 337.5 (185.2) | 250 | 321.3 (170.4) | 327.3 (183.7) | 5.6 (-25.3 to 36.6) | 0.72 | 0.6 (-30.3 to 31.4) | 0.97 |
| Alcohol consumption | 303 |  |  | 335 |  |  | 0.87 (0.65 to 1.18)<sup>d</sup> | 0.37 | 0.75 (0.54 to 1.04)<sup>e</sup> | 0.08 |
| Never |  | 117 (38.6%)<sup>c</sup> | 113 (37.3%)<sup>c</sup> |  | 126 (37.6%)<sup>c</sup> | 110 (32.8%)<sup>c</sup> |  |  |  |  |
| Monthly |  | 150 (49.5%)<sup>c</sup> | 151 (49.8%)<sup>c</sup> |  | 169 (50.4%)<sup>c</sup> | 183 (54.8%)<sup>c</sup> |  |  |  |  |
| Weekly |  | 36 (11.9%)<sup>c</sup> | 39 (12.9%)<sup>c</sup> |  | 40 (11.9%)<sup>c</sup> | 42 (12.5%)<sup>c</sup> |  |  |  |  |
| Physical activity (IPAQ-SF<br>categories) | 297 |  |  | 331 |  |  | **1.43 (1.06 to 1.91)<sup>d</sup>** | **0.02** | **1.41 (1.05 to 1.90)<sup>e</sup>** | **0.02** |
| Low activity |  | 119 (40.1%)<sup>c</sup> | 67 (22.6%)<sup>c</sup> |  | 129 (39.0%)<sup>c</sup> | 101 (30.5%)<sup>c</sup> |  |  |  |  |
| Moderate activity |  | 133 (44.8%)<sup>c</sup> | 146 (49.2%)<sup>c</sup> |  | 139 (42.0%)<sup>c</sup> | 155 (46.8%)<sup>c</sup> |  |  |  |  |
| High activity |  | 45 (15.2%)<sup>c</sup> | 84 (28.3%)<sup>c</sup> |  | 63 (19.0%)<sup>c</sup> | 75 (22.7%)<sup>c</sup> |  |  |  |  |
| **Outcomes at 6 months** |  |  |  |  |  |  |  |  |  |  |
| Diet (FFB scores) | 335 | 3.04 (0.4) | 3.17 (0.4) | 371 | 3.08 (0.4) | 3.14 (0.4) | **0.06 (0.01 to 0.11)** | **0.01** | **0.07 (0.02 to 0.11)** | **0.008** |
| Diet (sugary foods intake) | 333 | 2.25 (0.9) | 1.93 (0.8) | 367 | 2.18 (0.9) | 1.98 (0.9) | -0.10 (-0.20 to 0.00) | 0.05 | **-0.11 (-0.20 to -** | **0.04** |

|  | Intervention |  |  | Active Control |  |  | Diff in mean<sup>a</sup> | P | Adjusted<sup>b</sup> diff in<br>0.01) | P |
| --- | --- | --- | --- | --- | --- | --- | --- | --- | --- | --- |
| Physical Activity (IPAQ-SF MET minutes/ week) | 332 | 1462.9<br>(1449.4) | 1980.7<br>(2064.6) | 367 | 1516.9<br>(1545.0) | 1717.2<br>(1767.7) | **287.8**<br>**(21.1 to 554.4)** | **0.03** | 270.2<br>(3.1 to 537.4) | 0.05 |
| Sitting (IPAQ-SF minutes/ weekday) | 246 | 337.3 (183.8) | 325.2 (176.3) | 264 | 320.0 (167.8) | 316.1 (172.7) | 1.4 (-25.9 to 28.6) | 0.92 | 1.1 (-26.4 to 28.5) | 0.94 |
| Alcohol consumption | 335 |  |  | 371 |  |  | 1.01 (0.76 to 1.34)<sup>d</sup> | 0.97 | 0.91 (0.67 to 1.25)<sup>e</sup> | 0.57 |
| Never |  | 129 (38.5%)<sup>c</sup> | 129 (38.5%)<sup>c</sup> |  | 142 (38.3%)<sup>c</sup> | 145 (39.1%)<sup>c</sup> |  |  |  |  |
| Monthly |  | 168 (50.1%)<sup>c</sup> | 170 (50.7%)<sup>c</sup> |  | 187 (50.4%)<sup>c</sup> | 184 (49.6%)<sup>c</sup> |  |  |  |  |
| Weekly |  | 38 (11.3%)<sup>c</sup> | 36 (10.7%)<sup>c</sup> |  | 42 (11.3%)<sup>c</sup> | 42 (11.3%)<sup>c</sup> |  |  |  |  |
| Physical activity (IPAQ-SF categories) | 332 |  |  | 367 |  |  | **1.38 (1.05 to 1.83)<sup>d</sup>** | **0.02** | **1.35 (1.02 to 1.79)<sup>e</sup>** | **0.04** |
| Low activity |  | 132 (39.8%)<sup>c</sup> | 87 (26.2%)<sup>c</sup> |  | 149 (40.6%)<sup>c</sup> | 119 (32.4%)<sup>c</sup> |  |  |  |  |
| Moderate activity |  | 150 (45.2%)<sup>c</sup> | 153 (46.1%)<sup>c</sup> |  | 150 (40.9%)<sup>c</sup> | 170 (46.3%)<sup>c</sup> |  |  |  |  |
| High activity |  | 50 (15.1%)<sup>c</sup> | 92 (27.7%)<sup>c</sup> |  | 68 (18.5%)<sup>c</sup> | 78 (21.3%)<sup>c</sup> |  |  |  |  |

<sup>a</sup> Using ANCOVA (adjusting for baseline values).
<sup>b</sup> Same as <sup>a</sup> but additionally adjusting for site, recruitment pathway (NHS or community) and ethnicity (using categories shown in Table 1)
<sup>c</sup> Numbers and proportions with outcome at timepoint.
<sup>d</sup> Odds ratio for outcome at timepoint comparing intervention with active control.
<sup>e</sup> Odds ratio for outcome at timepoint comparing intervention with active control adjusting for site, recruitment pathway (NHS or community) and ethnicity (using categories shown in Table 1).

### Prespecified subgroup analyses

**Table 4:** Comparisons of weight (kg) between intervention and active control groups at 12 months by prespecified subgroups.

| Subgroup | Intervention |  |  | Active control |  |  | Diff in mean<sup>a</sup><br>(95% CI) | P interaction | Adjusted<sup>b</sup> diff in<br>mean (95% CI) | P interaction |
| --- | --- | --- | --- | --- | --- | --- | --- | --- | --- | --- |
|  | N | Baseline<br>Mean (SD) | Endpoint<br>Mean (SD) | N | Baseline<br>Mean (SD) | Endpoint<br>Mean (SD) |  |  |  |  |
| **Site** |  |  |  |  |  |  |  | 0.82 |  | 0.87 |
| Belfast | 69 | 84.4 (15.0) | 83.0 (14.6) | 84 | 86.7 (16.4) | 84.8 (17.3) | 0.4 (-1.7 to 2.5) |  | 0.6 (-1.5 to 2.7) |  |
| Bradford | 67 | 86.9 (16.5) | 85.3 (16.5) | 77 | 84.8 (18.4) | 83.7 (18.9) | -0.3 (-2.0 to 1.5) |  | -0.1 (-1.9 to 1.7) |  |
| London | 65 | 87.6 (16.7) | 85.7 (17.5) | 76 | 82.7 (15.7) | 81.8 (17.4) | -1.1 (-3.2 to 1.0) |  | -1.3 (-3.5 to 0.8) |  |
| Scotland | 62 | 87.3 (17.4) | 87.7 (20.3) | 58 | 91.5 (17.8) | 91.5 (18.9) | 0.6 (-1.4 to 2.7) |  | 0.6 (-1.5 to 2.7) |  |
| Cardiff | 60 | 85.8 (15.3) | 85.7 (15.3) | 56 | 87.2 (15.2) | 87.2 (17.6) | -0.1 (-2.1 to 1.9) |  | -0.3 (-2.4 to 1.8) |  |
| **Recruitment** |  |  |  |  |  |  |  | 0.13 |  | 0.12 |
| NHS | 20 | 88.9 (16.8) | 87.5 (15.6) | 18 | 95.2 (21.1) | 96.6 (22.2) | -3.0 (-6.6 to 0.5) |  | -3.4 (-7.6 to 0.8) |  |
| Community | 303 | 86.2 (16.1) | 85.3 (17.0) | 333 | 85.8 (16.6) | 84.8 (17.8) | 0.1 (-0.8 to 1.0) |  | 0.1 (-0.8 to 1.0) |  |
| **Ethnicity** |  |  |  |  |  |  |  | 0.26 |  | 0.27 |
| White | 228 | 87.3 (16.4) | 86.6 (17.2) | 224 | 88.0 (16.9) | 86.9 (18.2) | 0.3 (-0.8 to 1.4) |  | 0.2 (-0.9 to 1.3) |  |
| Mixed/Multiple/Other | 18 | 82.9 (14.1) | 79.9 (14.9) | 16 | 85.4 (16.5) | 85.3 (14.8) | -3.1 (-6.8 to 0.6) |  | -4.9 (-8.8 to -0.9) |  |
| Asian/Asian British | 43 | 78.7 (12.5) | 77.9 (12.9) | 69 | 78.8 (15.4) | 77.4 (16.5) | 0.7 (-1.1 to 2.5) |  | 0.3 (-1.6 to 2.1) |  |
| Black/Caribbean | 33 | 91.2 (16.4) | 89.9 (17.5) | 42 | 90.0 (16.6) | 90.4 (18.5) | -1.7 (-5.0 to 1.6) |  | -1.9 (-5.4 to 1.6) |  |
| **IMD** |  |  |  |  |  |  |  | 0.30 |  | 0.32 |
| 1<sup>st</sup> quintile- most deprived | 102 | 85.8 (15.2) | 85.0 (16.2) | 115 | 86.1 (18.9) | 85.0 (19.8) | 0.4 (-1.1 to 1.9) |  | 0.3 (-1.2 to 1.9) |  |
| 2<sup>nd</sup> quintile | 66 | 87.8 (17.4) | 86.1 (17.1) | 69 | 86.4 (17.5) | 86.4 (19.4) | -1.7 (-3.8 to 0.5) |  | -1.9 (-4.1 to 0.4) |  |
| 3<sup>rd</sup> quintile | 52 | 88.3 (16.1) | 87.3 (18.5) | 48 | 88.1 (14.5) | 88.0 (16.4) | -0.8 (-3.6 to 2.0) |  | -0.8 (-3.7 to 2.2) |  |
| 4<sup>th</sup> quintile | 41 | 89.3 (21.0) | 88.9 (21.5) | 61 | 87.4 (16.7) | 86.2 (17.6) | 0.7 (-1.2 to 2.7) |  | 0.6 (-1.4 to 2.6) |  |
| 5<sup>th</sup> quintile- least deprived | 62 | 82.3 (11.8) | 81.6 (12.0) | 58 | 83.8 (14.2) | 82.1 (14.9) | 1.0 (-1.0 to 3.0) |  | 0.8 (-1.3 to 3.0) |  |
| **Employment** |  |  |  |  |  |  |  | 0.23 |  | 0.24 |
| Full time | 133 | 86.0 (18.0) | 85.1 (18.5) | 161 | 87.1 (16.7) | 86.5 (18.1) | -0.4 (-1.8 to 1.1) |  | -0.5 (-2.0 to 1.0) |  |
| Part time | 107 | 86.0 (15.0) | 84.7 (15.7) | 109 | 82.0 (14.7) | 81.3 (15.0) | -0.5 (-1.8 to 0.8) |  | -0.4 (-1.8 to 0.9) |  |
| Unemployed/student | 77 | 87.7 (14.9) | 87.1 (15.9) | 78 | 90.0 (18.5) | 88.1 (20.7) | 1.5 (-0.5 to 3.5) |  | 1.4 (-0.6 to 3.4) |  |
| **Education (finished)** |  |  |  |  |  |  |  | 0.49 |  | 0.42 |

|  | Intervention |  |  | Active control |  |  | Diff in mean<sup>a</sup> | P interaction | Adjusted<sup>b</sup> diff in | P interaction |
| --- | --- | --- | --- | --- | --- | --- | --- | --- | --- | --- |
| Up to secondary (GCSE) | 26 | 89.1 (20.1) | 87.8 (21.4) | 34 | 87.1 (18.7) | 84.7 (20.9) | 1.1 (-2.4 to 4.7) |  | 0.8 (-2.6 to 4.3) |  |
| Secondary (A Level) | 30 | 84.3 (14.2) | 83.8 (14.2) | 31 | 93.8 (19.9) | 92.0 (20.1) | 0.9 (-1.9 to 3.7) |  | 0.8 (-2.3 to 3.9) |  |
| Further education | 45 | 86.8 (15.8) | 85.9 (16.3) | 45 | 89.7 (19.4) | 89.1 (20.8) | -0.3 (-3.0 to 2.3) |  | -0.5 (-3.4 to 2.4) |  |
| UG degree | 127 | 86.7 (16.8) | 85.5 (17.2) | 134 | 83.6 (14.2) | 83.4 (15.9) | -0.9 (-2.4 to 0.6) |  | -0.9 (-2.4 to 0.5) |  |
| PG degree | 95 | 85.7 (15.0) | 85.0 (16.5) | 107 | 85.8 (16.8) | 84.6 (17.9) | 0.4 (-1.1 to 1.9) |  | 0.6 (-0.9 to 2.2) |  |
| **Income** |  |  |  |  |  |  |  | 0.89 |  | 0.76 |
| Less than £30,000 | 105 | 87.7 (17.1) | 86.7 (18.1) | 104 | 87.3 (18.5) | 86.1 (20.1) | 0.2 (-1.5 to 1.9) |  | 0.4 (-1.3 to 2.2) |  |
| £30,001 or more | 197 | 85.7 (15.9) | 84.8 (16.4) | 222 | 86.2 (16.2) | 85.4 (17.4) | -0.1 (-1.2 to 1.0) |  | -0.2 (-1.4 to 0.9) |  |
| Don't know | 21 | 86.3 (13.8) | 84.8 (15.2) | 25 | 83.3 (15.8) | 82.4 (16.6) | -0.7 (-4.2 to 2.9) |  | -1.4 (-4.6 to 1.9) |  |
| **BMI (at study entry)** |  |  |  |  |  |  |  | 0.23 |  | 0.30 |
| 25.0-29.9 | 128 | 74.0 (6.9) | 73.6 (8.2) | 146 | 73.9 (6.8) | 72.8 (8.4) | 0.6 (-0.5 to 1.8) |  | 0.6 (-0.5 to 1.8) |  |
| 30.0 and over | 195 | 94.5 (15.3) | 93.2 (16.7) | 205 | 95.1 (16.4) | 94.3 (18.0) | -0.5 (-1.8 to 0.8) |  | -0.5 (-1.8 to 0.8) |  |
| **Parity** |  |  |  |  |  |  |  | 0.33 |  | 0.40 |
| 1 | 137 | 86.0 (15.8) | 85.8 (16.4) | 150 | 85.5 (15.9) | 84.6 (17.4) | 0.6 (-0.7 to 1.9) |  | 0.7 (-0.6 to 2.0) |  |
| 2 | 124 | 86.2 (17.3) | 85.3 (17.9) | 114 | 87.5 (17.9) | 86.8 (19.0) | -0.3 (-1.8 to 1.2) |  | -0.3 (-1.8 to 1.3) |  |
| 3+ | 62 | 87.5 (14.6) | 84.9 (16.1) | 87 | 86.2 (17.3) | 84.8 (18.4) | -1.2 (-3.3 to 1.0) |  | -0.7 (-2.9 to 1.5) |  |
| **Weeks postpartum at study entry** |  |  |  |  |  |  |  | 0.04 |  | 0.12 |
| ≤26 weeks (≤6 months) | 121 | 86.8 (16.6) | 85.4 (18.0) | 120 | 85.6 (16.8) | 83.5 (18.0) | 0.7 (-0.9 to 2.2) |  | 0.4 (-1.2 to 1.9) |  |
| 27-52 weeks (7-12 months) | 88 | 85.7 (15.1) | 85.3 (15.9) | 103 | 87.6 (17.5) | 86.1 (18.4) | 1.0 (-0.6 to 2.7) |  | 1.0 (-0.7 to 2.7) |  |
| ≥53 weeks (>12 months) | 114 | 86.5 (16.6) | 85.6 (16.6) | 128 | 85.9 (16.6) | 86.6 (18.1) | -1.5 (-2.9 to -0.1) |  | -1.4 (-2.9 to 0.0) |  |

<sup>a</sup> Using ANCOVA (adjusting for baseline values).
<sup>b</sup> Same as <sup>a</sup> but additionally adjusting for site, recruitment pathway (NHS or community) and ethnicity (using categories shown in Table 1).

### Acceptability of trial methods

Findings relevant to the acceptability of trial methods are summarised in the Supplementary information and Supplementary Tables 6 and 7.

### Fidelity of trial blinding

Three instances of researcher unblinding were recorded as follows: 1) an email sent by the LSHTM text message platform manager to a site researcher while resolving a data entry error in the LSHTM randomisation system revealed the allocation of two participants; 2) a text message sent by a participant to a site researcher revealed group allocation; and, 3) an email sent by a participant to the main trial email address revealed group allocation.

### Adverse events

There were 75 SAEs recorded across the trial (April 2022-June 2024). None were categorised as severe, related and unexpected and there was no onward reporting of SAEs required.

## Discussion

### Statement of principal findings

The trial met its recruitment and retention targets and recruited an ethnically and socioeconomically diverse sample of women from across the UK. In this two-arm, parallel group RCT of a 12-month automated text message-delivered weight management intervention, weight at 12 months was not meaningfully reduced for postpartum women with overweight or obesity allocated to the intervention group compared with the active control. There were small improvements in the secondary endpoints, diet, physical activity and infant feeding behaviours, for the intervention group compared to active control at 12 months, including a shift from low to moderate/high physical activity levels. However, the reduced consumption of sugar-rich foods observed at 6 months in the intervention group relative to the control was not sustained at 12 months. We observed high participant retention and acceptability of the trial methods across the diverse sample of participants.

### Strengths and limitations of the study

The SMS intervention is theoretically informed, developed with PPI and previously pilot tested for feasibility and acceptability, then culturally adapted for a diverse UK population prior to the full effectiveness trial (35, 36). The robust trial delivery provides internal and external validity in our findings, tackling persistent challenges in recruitment and retention encountered in postpartum weight management trials (10). The methods, specifically designed for the target population and flexibly tailored across study localities, were highly acceptable and target recruitment and high retention rates were achieved across all sub-groups. We achieved adequate power to examine the long-term primary outcome. The participants recruited were diverse and representative of the UK population in terms of ethnicity, IMD and geography across all four UK countries. This allowed examination of the effect across different subgroups of women ensuring generalisability of our findings and simultaneously indicating no inadvertent widening of inequalities because of the intervention. While weight and BMI outcomes alone do not fully capture health risk, we prioritised offering home visits and minimising participant burden which is critical for this population, and therefore limited body composition measures to waist circumference rather than more invasive and time-consuming assessments.

### Strengths and weaknesses in relation to other studies, discussing important differences in results

Women have previously shown willingness to participate in postpartum behavioural weight management interventions (50). However, the approaches taken have often been resource-intensive and failed to consider the population specific barriers to participation and engagement (10, 19, 20, 51). The SMS trial aimed to provide an accessible, sustainable and scalable solution to supporting postpartum weight management. It addressed many evidence gaps (52) in a way that was sensitive to the needs of postpartum women, offering flexible trial methods that enabled participation by reducing the inconvenience and cost of travel, and accounting for childcare commitments and lack of time to devote to intensive in-person interventions (19, 51).

Most postpartum trials recruit women during pregnancy or shortly after birth, with some extending to 12 (46, 53–58) and 18 months (59) postpartum. By recruiting up to 24 months postpartum, we aimed to address the limited evidence on when to begin postpartum weight management support (5, 10, 60) and to align with relevant NICE guidance (8) on supporting postpartum weight management. We enrolled women across this extended postpartum period, indicating both sustained interest in support and wide variation in readiness to engage. These findings highlight the value of offering flexible initiation windows in future postpartum behavioural weight management interventions. We will further explore optimal timing in the trial process evaluation (see the PEAP in Supplementary information).

Systematic review evidence indicates that using communication technologies to support weight reduction among postpartum women is beneficial (22, 23, 52). However, a 2019 review identified just eight RCTs using technology for postpartum weight loss, seven of which used it as a supplement to in-person counselling rather than the sole mode of delivery (22). The literature is also limited by small sample sizes and short follow-up periods, with most trials having between 18-66 participants and follow-up lasting between 40 days and 16 □weeks. Trials of behavioural weight management interventions including technology-based approaches for postpartum women with overweight or obesity published since this review have mainly been pilot or feasibility trials, have used technology alongside other approaches, and many have continued to experience recruitment and retention difficulties that, for some, have been further compounded by the impact of COVID-19 lockdowns (53, 61–65). To our knowledge, there are currently no other published RCTs of behavioural weight management interventions delivered solely by fully automated text messages to support postpartum weight management for women with overweight or obesity. In delivering an adequately powered trial, with both intervention duration and the assessment of outcomes over a long-term 12 month period, we have strengthened the quality of the existing evidence and addressed a gap in determining the independent effects of this technology on postpartum weight management (23, 52).

Two comparable, adequately powered, 12 month trials have examined the effectiveness of mobile-health (m-Health) behavioural interventions for postpartum women with overweight or obesity (46, 65). The 12-month SnapBack intervention used text messages alongside coaching telephone calls and Facebook support, to promote postpartum weight loss compared with usual care for socioeconomically disadvantaged African American women (n= 300) attending Women, Infants and Children (WIC) clinics (65). The SnapBack intervention did not result in weight loss at 6 or 12 months. There was a beneficial effect of the intervention on systolic blood pressure, but dietary and physical activity behaviours were not reported. On the other hand, a 12 month internet-based intervention approach to support postpartum weight loss in low-income postpartum women (n= 371) attending WIC clinics supported a reduction in weight at 12 months compared with the standard care control (mean difference: 2.3 kg (95% CI, 1.1 to 3.5), *p* < .001) but no substantial changes in dietary and physical activity outcomes (46). Although predominantly m-Health delivered, both interventions included other resource intensive components such as in-person groups and coaching, so it is difficult to distinguish the impacts attributable to the technology-based aspects alone. There is also uncertainty about the effects of these interventions for different groups of women.

Some recent feasibility trials of m-Health delivered postpartum behavioural weight management interventions have shown significant reductions in weight outcomes (53, 64) but are limited by small study samples (n= 22/62) with a lack of racial, ethnic and socioeconomic diversity, and shorter intervention durations (14 weeks/6 months). In our trial, the intervention did support small, positive, sustained impacts on diet, physical activity and infant feeding at 12 months and a notable shift from low to moderate or high activity levels. Previous postpartum studies that have examined these outcomes have shown varied results in relation to diet and activity behaviour changes (20, 50, 54, 61, 63) and have mainly been feasibility studies with shorter intervention durations. We identified one trial similarly showing sustained improvements in diet outcomes at 12 months (66).

### Meaning of the study: possible explanations and implications for clinicians and policymakers

Viewed alongside the existing evidence, this trial demonstrates the considerable challenge that all mothers face in achieving clinically meaningful weight loss after childbirth despite encouraging positive changes in health-promoting behaviours noted throughout the intervention. Although the SMS intervention was designed with cognisance of this challenge, our findings suggest that there is no universally appropriate approach to postpartum weight management that is likely to work for all women. There may be merit in exploring the potential use of data-driven models, such as machine learning or artificial intelligence, alongside behavioural theories, to develop more accurate and adaptable models of behaviour which allow for more dynamic tailoring of interventions to maximise engagement (72, 73).

Nevertheless, given the positive findings in relation to diet, activity and infant feeding behaviours, this intervention could help fill a gap in the support currently offered in the postpartum period, whilst also being complementary to other weight management strategies for those needing more intensive approaches. Of note is the continued need for scalable behavioural diet, physical activity and weight management interventions, such as SMS, alongside the growing use of weight loss medications to treat obesity. Recent guidance from the NICE and the World Health Organisation recommend wraparound care, including diet, nutrition and physical activity support, for people using weight loss medications, as well as continued behavioural therapy after stopping them (74, 75). In the context of our trial population, Glucagon-Like Peptide-1 medications are unsuitable while trying to conceive, during pregnancy or while breastfeeding (76). Holistic interventions that include promotion of sustained behaviour change therefore remain essential for long-term weight management and improved health for all.

### Unanswered questions and future research

In addition to offering a more accessible delivery mode for postpartum women, text message-based interventions are proposed to offer a low-cost and scalable solution for supporting weight management in this population, without added burden to an over-stretched National Health Service. However, cost-effectiveness of such interventions has yet to be examined.

The planned cost-effectiveness evaluation of the SMS intervention will provide further important evidence on the potential of using this text message-based approach.

The planned, mixed-methods process evaluation will fully explore how women respond to and interact with the intervention and its acceptability, alongside examining implementation, the wider benefits or impacts of receiving the intervention and the active control, including interactions with mental health and any other contextual factors influencing outcomes (see the PEAP in Supplementary information). Furthermore, we will examine the effect of the intervention on theoretical mediators of change, such as self-esteem and self-efficacy, that, alongside weight and health behaviours, are important women-centred outcomes to consider.

Few trials of postpartum behavioural weight management interventions for women with overweight or obesity have examined the longer-term postpartum weight trends or impacts of intervention participation beyond 18 months postpartum (5, 77, 78). We will examine outcomes for women in the SMS trial at 24 months to help address this evidence gap and to further understand postpartum weight trajectories across this ethnically and socioeconomically diverse UK population.

## Conclusion

The SMS trial makes a novel and important contribution to the evidence on effectiveness of using automated text messages to support postpartum weight management. In an ethnically and socioeconomically diverse sample of UK postpartum women with overweight or obesity, the 12-month behavioural intervention did not lead to meaningful weight loss compared with an active control group receiving child health and development messages, but it did support small, sustained improvements in diet, physical activity and infant feeding behaviours.

## Supporting information

Code used for data analysis

## Contributor information

MCM was the Principal Investigator. Concept and design - DG, SB, EC, CFr, PH, ASA, CRC, SUD, SH, FK, CM, EM, JVW and MCM. Acquisition, analysis, or interpretation of data - DG, ES, NC-M, KC, CFe, AI, BK, CBK, ML, EO-A, RP, HS, SB, EC, CFr, PH, ASA, CRC, SUD, SH, FK, CM, EM, LM, JVW and MCM; Public contributor – SH; Writing - original draft preparation – DG, ES and MCM; Writing - review and editing – DG, ES, NC- M, KC, CFe, AI, BK, CBK, ML, EO-A, RP, HS, SB, EC, CFr, PH, ASA, CRC, SUD, SH, FK, CM, EM, LM, JVW and MCM; Supervision - SB, EC, CFr, PH and MCM; Administration – DG, SB, EC, CFr, PH and MCM; Funding acquisition - DG, SB, EC, CFr, PH, ASA, CRC, SUD, FK, CM, EM, JVW and MCM. All authors (DG, ES, NC-M, KC, CFe, AI, BK, CBK, ML, EO-A, RP, HS, SB, EC, CFr, PH, ASA, CRC, SUD, SH, FK, CM, EM, LM, JVW and MCM) contributed to the revision of the manuscript and have read and approved the final version. The corresponding author attests that all listed authors meet authorship criteria and that no others meeting the criteria have been omitted.

The authors would like to acknowledge the valuable inputs made by all public contributors who informed the design and delivery of the study. The authors would also like to thank all the women who took part.

## Funding and statement of researcher independence

This work was funded by the National Institute for Health and Care Research (NIHR) Public Health Research Programme (NIHR131509) (https://fundingawards.nihr.ac.uk/award/NIHR131509) undergoing external peer review at application. The funder had no role in the study design, data collection, analysis and interpretation, or in the writing of this manuscript. Intervention costs were provided by the HSC Research and Development Division of the Public Health Agency Northern Ireland.

The trial sponsor is Queen’s University Belfast. The sponsor was not responsible for the trial design, collection, management, analysis or interpretation of data or report writing but had oversight of trial progress, including publications.

## Ethical approval

Ethical approval was obtained from the West of Scotland Research Ethics Service REC 4 22/WS/0003 (IRAS: 305557). The study adhered to the Declaration of Helsinki and written informed consent was collected from all participants.

## Data sharing statement

The supplementary information provides the code used for data analysis in this study. The data underpinning the presented findings are openly and publicly accessible (https://doi.org/10.5061/dryad.pg4f4qs5b). If you encounter problems accessing the data, please contact the corresponding author.

## Transparency statement

The lead author affirms that the manuscript is an honest, accurate, and transparent account of the study being reported; that no important aspects of the study have been omitted; and that any discrepancies from the study as originally planned have been explained.

## Dissemination to participants and related patient and public communities

Once the results are published, the findings of this study will be disseminated to participants using multiple formats based on participants’ preferences (see PPI in methods). We will disseminate the results to the wider public, and participating sites and organisations, through our university websites and social media. Findings will be shared with researchers, clinicians and policymakers/public health bodies through conferences and a policy briefing.

## Provenance and peer review

Not commissioned; externally peer reviewed.

## Copyright/license for publication

For the purpose of open access, a Creative Commons Attribution (CC BY) licence is required. The Corresponding Author has the right to grant on behalf of all authors and does grant on behalf of all authors, a worldwide licence to the Publishers and its licensees in perpetuity, in all forms, formats and media (whether known now or created in the future), to i) publish, reproduce, distribute, display and store the Contribution, ii) translate the Contribution into other languages, create adaptations, reprints, include within collections and create summaries, extracts and/or, abstracts of the Contribution, iii) create any other derivative work(s) based on the Contribution, iv) to exploit all subsidiary rights in the Contribution, v) the inclusion of electronic links from the Contribution to third party material where-ever it may be located; and, vi) licence any third party to do any or all of the above.

## Data Availability

The supplementary information provides the code used for data analysis in this study. All data produced are openly and publicly accessible online at https://doi.org/10.5061/dryad.pg4f4qs5b.

