## Supplementary Figure 1 for "Effectiveness of an automated text message intervention for weight management in postpartum women with overweight or obesity (Supporting MumS (SMS)): a UK wide, multicentre, two arm, parallel group, randomised controlledti trial"

**Supplementary Figure 1. Forest plot of adjusted mean difference in weight (and 95% confidence intervals) between intervention and active control group at 12 months by site.**

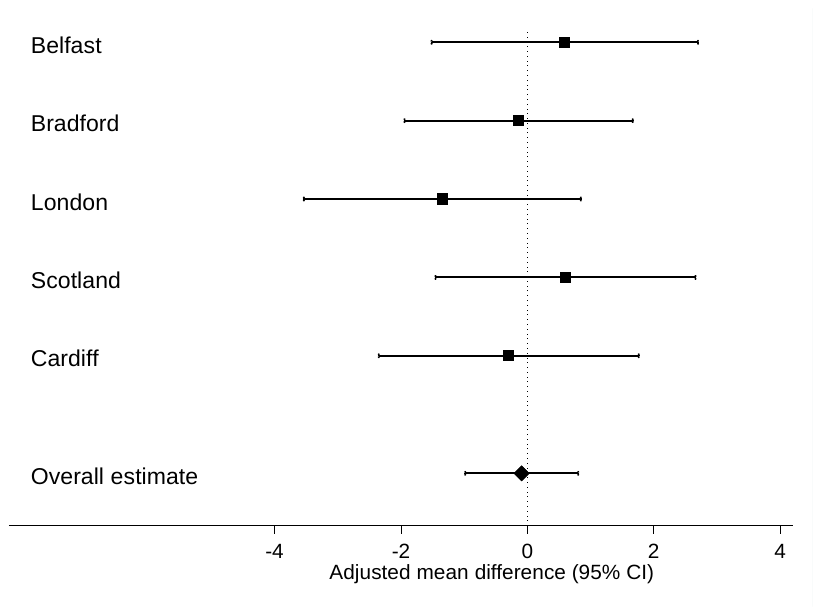
