## Supplementary material for "Effectiveness of an automated text message intervention for weight management in postpartum women with overweight or obesity (Supporting MumS (SMS)): a UK wide, multicentre, two arm, parallel group, randomised controlledti trial": Code used for data analysis

log using "F:\Smums\Final analysis\Log file after BMJ code.log", replace

*FINAL BMJ 5th October 2026.
import spss using "F:\Smums\Final analysis\Primary paper dataset_V3.0_190626.sav", clear 

*Sort on patient id (ensures imputation results match paper).
sort PID

*Change all variable names to lower case.
foreach var of varlist * {
  rename `var' `=strlower("`var'")'
}

*Create variable that is recorded weight at 12 months expcept if recorded weight is missing and then it is self recorded weight.
gen     m12_wghtplusself= m12_wght_kg
replace m12_wghtplusself= m12_srwght_kg_rec if m12_wghtplusself==.

*Recode weight at 12 months to missing if weight loss medications or surgery reported at 6 or 12 months.
gen     m12_wghtnomeds=m12_wght_kg 
replace m12_wghtnomeds=. if m12_weightlossmeds==1 | m12_weightlosssurgery==1 | m6_weightlossmeds==1 | m6_weightlosssurgery==1

*Create a variable that recodes the 12 month weight as the baseline weight if the 12 month weight is missing.
gen     m12_wghtbocf=m12_wght_kg
replace m12_wghtbocf=bl_wght_kg if  m12_wghtbocf==.

*Create a variable that recodes the 12 month weight to the 6 month weight if the 12 month weight is missing and recodes the 12 month weight to the baseline weight if the 12 and 6 month weight are missing.
gen     m12_wghtlocf=m12_wght_kg
replace m12_wghtlocf=m6_wght_kg if  m12_wghtlocf==.
replace m12_wghtlocf=bl_wght_kg if  m12_wghtlocf==.

*Creat a variable that captures and weight loss medications or surgery at 6 or 12 months.
gen medssurg12=1 if  m12_weightlossmeds==1 | m12_weightlosssurgery==1 | m6_weightlossmeds==1 | m6_weightlosssurgery==1
replace medssurg12=0 if medssurg12==.

*Recode ethnicity group.
replace bl_ethnorg=bl_ethnicgroup_rec
replace bl_ethnorg=. if bl_ethnorg==-9

*Recode treatment group.
gen treatment=group
replace treatment=0 if group==2

*Recode parity.
replace bl_parity=3 if bl_parity>=3 & bl_parity<7
label define number 1 1 2 2 3 "3+" 
label values bl_parity number

*Add categories names to stage post partum.
rename  stageppcategories blpostpart
label define blpostpart 1 "<=26" 2 "27-52" 3 ">52" 
label values blpostpart blpostpart

*Recodes bmi to missing if bmi based upon selfreported weight.
replace m12_bmi=. if m12_wght_kg==. & m12_bmi~=.
replace m6_bmi=. if m6_wght_kg==. & m6_bmi~=.

*Rename employment and add category names.
gen bl_empl_recode=1 if bl_employ==1
replace bl_empl_recode=2 if bl_employ==2
replace bl_empl_recode=3 if bl_employ==3 
label define bl_empl_recode 1 "FT" 2 "PT" 3 "Unemploy/student"
label values bl_empl_recode bl_empl_recode

*Recode marital status.
gen bl_mstat_recode=1 if bl_mstatus==1 | bl_mstatus==5
replace bl_mstat_recode=2 if bl_mstatus==2 |  bl_mstatus==3 | bl_mstatus==4
replace bl_mstat_recode=3 if bl_mstatus==3
label define bl_mstat_recode 1 "Sing+sep" 2 "Married/cvilp/livingwihtp" 3 "Other" 
label values bl_mstat_recode bl_mstat_recode

*Rename baseline variables.
gen bl_ffbscore=m0_ffbscore
gen bl_totalpa_metmins=m0_totalpa_metmins
gen bl_meansugar=m0_meansugar
gen bl_totalsitting=m0_totalsitting

save "F:\Smums\Final analysis\bmjfinalsmumsdata",replace


*Table 1.
use "F:\Smums\Final analysis\bmjfinalsmumsdata",clear

*Check dustribution of continuous baseline variables.
histogram bl_age
histogram bl_height
histogram bl_wght_kg
histogram bl_bmi
histogram bl_waistcir

*Calculate summary statistics for continuous variables by intervention group in participants as randomised.
summarize bl_age if treatment==0 
summarize bl_age if treatment==1 
summarize bl_height if treatment==0 
summarize bl_height if treatment==1 
summarize bl_wght_kg if treatment==0 
summarize bl_wght_kg if treatment==1 
summarize bl_bmi if treatment==0 
summarize bl_bmi if treatment==1 
summarize bl_waistcir if treatment==0 
summarize bl_waistcir if treatment==1 
*Calculate frequency tables for categorical variables by intervention group in participants as randomised.
tab  bl_bmi_categories treatment ,col
tab site treatment  ,col
tab recruitmentpathway_subgroups treatment ,col
tab  bl_ethnorg treatment ,col
tab  imd_quintile treatment  ,col
tab  bl_employ treatment  ,col
tab  bl_education treatment  ,col
tab  bl_income treatment  ,col
tab  bl_mstatus treatment  ,col
tab  bl_parity treatment  ,col
tab  bl_alcohol treatment  ,col
tab  bl_smok treatment  ,col
tab  blpostpart treatment  ,col
tab  bl_epdsabove9 treatment  ,col
tab  bl_hthcond treatment  ,col

*Calculate summary statistics for continuous variables by intervention group, restricted to particpants who contributed to main analysis.
summarize bl_age if treatment==0 & m12_wght_kg~=. & bl_wght_kg~=. 
summarize bl_age if treatment==1 & m12_wght_kg~=. & bl_wght_kg~=.
summarize bl_height if treatment==0 & m12_wght_kg~=. & bl_wght_kg~=. 
summarize bl_height if treatment==1 & m12_wght_kg~=. & bl_wght_kg~=.
summarize bl_wght_kg if treatment==0 & m12_wght_kg~=. & bl_wght_kg~=.
summarize bl_wght_kg if treatment==1 & m12_wght_kg~=. & bl_wght_kg~=.
summarize bl_bmi if treatment==0 & m12_wght_kg~=. & bl_wght_kg~=.
summarize bl_bmi if treatment==1 & m12_wght_kg~=. & bl_wght_kg~=.
summarize bl_waistcir if treatment==0 & m12_wght_kg~=. & bl_wght_kg~=.
summarize bl_waistcir if treatment==1 & m12_wght_kg~=. & bl_wght_kg~=.
*Calculate frequency tables for categorical variables by intervention group, restricted to particpants who contributed to main analysis.
tab  bl_bmi_categories treatment if m12_wght_kg~=. & bl_wght_kg~=.,col
tab site treatment if m12_wght_kg~=. & bl_wght_kg~=. ,col
tab recruitmentpathway_subgroups treatment if m12_wght_kg~=. & bl_wght_kg~=.,col
tab  bl_ethnorg treatment if m12_wght_kg~=. & bl_wght_kg~=.,col
tab  imd_quintile treatment if m12_wght_kg~=. & bl_wght_kg~=. ,col
tab  bl_employ treatment if m12_wght_kg~=. & bl_wght_kg~=. ,col
tab  bl_education treatment if m12_wght_kg~=. & bl_wght_kg~=. ,col
tab  bl_income treatment if m12_wght_kg~=. & bl_wght_kg~=. ,col
tab  bl_mstatus treatment if m12_wght_kg~=. & bl_wght_kg~=. ,col
tab  bl_parity treatment if m12_wght_kg~=. & bl_wght_kg~=. ,col
tab  bl_alcohol treatment if m12_wght_kg~=. & bl_wght_kg~=. ,col
tab  bl_smok treatment if m12_wght_kg~=. & bl_wght_kg~=. ,col
tab  blpostpart treatment if m12_wght_kg~=. & bl_wght_kg~=. ,col
tab  bl_epdsabove9 treatment if m12_wght_kg~=. & bl_wght_kg~=. ,col
tab  bl_hthcond treatment if m12_wght_kg~=. & bl_wght_kg~=. ,col

*Table 2: Comparisons of weight and body composition outcomes between intervention and active control at 6 and 12 months, including sensitivity analysis.
use "F:\Smums\Final analysis\bmjfinalsmumsdata",clear

*Analyses at 12 months.
*Weight, waist circumference and BMI.
foreach j in wght_kg waistcir bmi{
*Calculate baseline mean and standard deviation by intervention group, restricted to participants with 12 month and baseline value.
summarize  bl_`j' if treatment==0 & m12_`j'~=. & bl_`j'~=. 
summarize  bl_`j' if treatment==1 & m12_`j'~=. & bl_`j'~=. 
*Calculate 12 month mean and standard deviation by intervention group, restricted to participants with 12 month and baseline value.
summarize  m12_`j' if treatment==0 & m12_`j'~=. & bl_`j'~=. 
summarize  m12_`j' if treatment==1 & m12_`j'~=. & bl_`j'~=. 
*Conduct ANCOVA to calculate difference in mean by intervention group (and 95% CI and P value) adjusting for baseline.
regress  m12_`j' i.treatment bl_`j'
*Conduct ANCOVA to calculate difference in mean by intervention group (and 95% CI and P value) adjusting for baseline and site, recruitment pathway and ethnicity. 
regress  m12_`j' i.treatment i.site i.recruitmentpathway_subgroups i.bl_ethnorg bl_`j'
*Generate residuals and check model fit.
predict residm12_`j',resid
predict fitm12_`j',xb
histogram residm12_`j'
scatter residm12_`j' fitm12_`j'
}

*Analyses at 12 months.
*Weight gain >5kg.
use "F:\Smums\Final analysis\bmjfinalsmumsdata",clear
*Calculate weight gain greater than 5kg at 12 months.
gen gain=m12_wght_kg-bl_wght_kg
gen gain5kg=1 if gain>=5 & gain~=.
replace gain5kg=0 if gain<5 & gain~=.
*Count number with weight gain greater than 5kg bu intervention group.
count if gain5kg~=. & treatment==0
count if gain5kg~=. & treatment==1
count if gain5kg==1 & treatment==0
count if gain5kg==1 & treatment==1
*Conduct logistic regression to calculate OR for weight gain by intervention group (and 95% CI and P value).
logit gain5kg i.treatment,or
*Conduct logistic regression to calculate OR for weight gain by intervention group (and 95% CI and P value) adjusting for site, recruitment pathway and ethnicity.
logit  gain5kg i.treatment i.site i.recruitmentpathway_subgroups i.bl_ethnorg,or

*Analyses at 6 months
*Weight, waist circumference and BMI
use "F:\Smums\Final analysis\bmjfinalsmumsdata",clear
foreach j in wght_kg waistcir bmi{
*Calculate baseline mean and standard deviation by intervention group, restricted to participants with 6 month and baseline value.
summarize  bl_`j' if treatment==0 & m6_`j'~=. & bl_`j'~=. 
summarize  bl_`j' if treatment==1 & m6_`j'~=. & bl_`j'~=. 
*Calculate 6 month mean and standard deviation by intervention group, restricted to participants with 6 month and baseline value.
summarize  m6_`j' if treatment==0 & m6_`j'~=. & bl_`j'~=. 
summarize  m6_`j' if treatment==1 & m6_`j'~=. & bl_`j'~=. 
*Conduct ANCOVA to calculate difference in mean by intervention group (and 95% CI and P value) adjusting for baseline.
regress  m6_`j' i.treatment bl_`j'
*Conduct ANCOVA to calculate difference in mean by intervention group (and 95% CI and P value) adjusting for baseline and site, recruitment pathway and ethnicity. 
regress  m6_`j' i.treatment i.site i.recruitmentpathway_subgroups i.bl_ethnorg bl_`j'
*Generate residuals and check model fit.
predict residm6_`j',resid
predict fitm6_`j',xb
histogram residm6_`j'
scatter residm6_`j' fitm6_`j'
}

*Analyses at 6 months
*Weight gain >5kg
use "F:\Smums\Final analysis\bmjfinalsmumsdata",clear
*Calculate weight gain greater than 5kg at 6 months.
gen gain=m6_wght_kg-bl_wght_kg
gen gain5kg=1 if gain>=5 & gain~=.
replace gain5kg=0 if gain<5 & gain~=.
*Count number with weight gain greater than 5kg bu intervention group.
count if gain5kg~=. & treatment==0
count if gain5kg~=. & treatment==1
count if gain5kg==1 & treatment==0
count if gain5kg==1 & treatment==1
*Conduct logistic regression to calculate OR for weight gain by intervention group (and 95% CI and P value).
logit gain5kg i.treatment,or
logit  gain5kg i.treatment i.site i.recruitmentpathway_subgroups i.bl_ethnorg,or

*Sensitivity analyses
*Measured weight including self reported (m12_wghtplusself), weight excluding participants using weight loss meds/surgery (m12_wghtnomeds), baseline observation carried forward (bocf), last observation carried forward (locf)  
foreach j in m12_wghtplusself m12_wghtnomeds m12_wghtbocf m12_wghtlocf{
use "F:\Smums\Final analysis\bmjfinalsmumsdata",clear
summarize  bl_wght_kg if treatment==0 & `j'~=. & bl_wght_kg~=. 
summarize  bl_wght_kg if treatment==1 & `j'~=. & bl_wght_kg~=. 
summarize  `j' if treatment==0 & `j'~=. & bl_wght_kg~=. 
summarize  `j' if treatment==1 & `j'~=. & bl_wght_kg~=. 
*Conduct ANCOVA to calculate difference in mean by intervention group (and 95% CI and P value) adjusting for baseline.
regress  `j' i.treatment bl_wght_kg
*Conduct ANCOVA to calculate difference in mean by intervention group (and 95% CI and P value) adjusting for baseline and site, recruitment pathway and ethnicity. 
regress  `j' i.treatment i.site i.recruitmentpathway_subgroups i.bl_ethnorg bl_wght_kg
predict residm12_`j',resid
predict fitm12_`j',xb
histogram residm12_`j'
scatter residm12_`j' fitm12_`j'
}

*Weight multiple imputation
use "F:\Smums\Final analysis\bmjfinalsmumsdata",clear
mi set flong
mi register imputed m12_wght_kg
*Impute missing weight at 12 months using treatment, site, recruitment pathway, ethnicity and baseline weight. 
mi impute regress m12_wght_kg = i.treatment i.site i.recruitmentpathway_subgroups i.bl_ethnorg bl_wght_kg ,add(10) rseed(1000) noisily
summarize  bl_wght_kg if treatment==0 & m12_wght_kg~=. & bl_wght_kg~=. & _mi_m==1
summarize  bl_wght_kg if treatment==1 & m12_wght_kg~=. & bl_wght_kg~=.  & _mi_m==1 
summarize  m12_wght_kg if treatment==0 & m12_wght_kg~=. & bl_wght_kg~=.  & _mi_m==1
summarize  m12_wght_kg if treatment==1 & m12_wght_kg~=. & bl_wght_kg~=.  & _mi_m==1
*Conduct ANCOVA to calculate difference in mean by intervention group (and 95% CI and P value) adjusting for baseline and pool across imputed datasets.
mi estimate:regress  m12_wght_kg i.treatment bl_wght_kg
*Conduct ANCOVA to calculate difference in mean by intervention group (and 95% CI and P value) adjusting for baseline and site, recruitment pathway and ethnicity and pool across imputed datasets. 
mi estimate:regress  m12_wght_kg i.treatment bl_wght_kg i.site i.recruitmentpathway_subgroups i.bl_ethnorg 

*Weight multiple imputation using delta method
use "F:\Smums\Final analysis\bmjfinalsmumsdata",clear
mi set flong
mi register imputed m12_wght_kg
mi impute regress m12_wght_kg = i.treatment i.site i.recruitmentpathway_subgroups i.bl_ethnorg bl_wght_kg ,add(10) rseed(1000) noisily
*Impute missing weight at 12 months using treatment, site, recruitment pathway, ethnicity and baseline weight. 
mi passive: generate byte imputed=_mi_miss
replace imputed =1 if imputed==.
generate float m12_wght_kg_delta=m12_wght_kg
*Increase imputed weights by an average of 0.91 kgs.
replace        m12_wght_kg_delta=m12_wght_kg+0.91 if imputed==1
summarize  bl_wght_kg if treatment==0 & m12_wght_kg~=. & bl_wght_kg~=. & _mi_m==1
summarize  bl_wght_kg if treatment==1 & m12_wght_kg~=. & bl_wght_kg~=.  & _mi_m==1 
summarize  m12_wght_kg_delta if treatment==0 & m12_wght_kg~=. & bl_wght_kg~=.  & _mi_m==1
summarize  m12_wght_kg_delta if treatment==1 & m12_wght_kg~=. & bl_wght_kg~=.  & _mi_m==1
*Conduct ANCOVA to calculate difference in mean by intervention group (and 95% CI and P value) adjusting for baseline and pool across imputed datasets.
mi estimate:regress  m12_wght_kg_delta i.treatment bl_wght_kg
*Conduct ANCOVA to calculate difference in mean by intervention group (and 95% CI and P value) adjusting for baseline and site, recruitment pathway and ethnicity and pool across imputed datasets. 
mi estimate:regress  m12_wght_kg_delta i.treatment bl_wght_kg i.site i.recruitmentpathway_subgroups i.bl_ethnorg 

**Table 3: Comparisons of secondary outcomes between intervention and active control at 6 and 12 months
*Continuous outcomes: ffb score, sugary food intake, physical activity and sitting time
clear
use "F:\Smums\Final analysis\bmjfinalsmumsdata",clear

*Conduct analysis for 12 month and 6 month outcomes.
foreach z in 12 6 {
	*Conduct analysis for Diet-FFB score (ffbscore), Diet-Sugar food intake (meansugar), Physical activity (totalpa_metmins) and Sitting time (totalsitting). 
	foreach j in ffbscore meansugar totalpa_metmins totalsitting {
	*Calculate baseline mean and standard deviation by intervention group, restricted to participants with 12/6 month and baseline value.
	summarize  m0_`j' if treatment==0 & m`z'_`j'~=. & m0_`j'~=. 
	summarize  m0_`j' if treatment==1 & m`z'_`j'~=. & m0_`j'~=. 
	*Calculate 12/6 month mean and standard deviation by intervention group, restricted to participants with 12/6 month and baseline value.
	summarize  m`z'_`j' if treatment==0 & m`z'_`j'~=. & m0_`j'~=. 
	summarize  m`z'_`j' if treatment==1 & m`z'_`j'~=. & m0_`j'~=. 
	*Conduct ANCOVA to calculate difference in mean by intervention group (and 95% CI and P value) adjusting for baseline.
	regress  m`z'_`j' i.treatment m0_`j'
	*Conduct ANCOVA to calculate difference in mean by intervention group (and 95% CI and P value) adjusting for baseline and site, recruitment pathway and ethnicity. 
	regress  m`z'_`j' i.treatment i.site i.recruitmentpathway_subgroups i.bl_ethnorg m0_`j'
	*Generate residuals and check model fit.
	predict resid`z'_`j',resid
	predict fitm`z'_`j',xb
	histogram resid`z'_`j'
	scatter resid`z'_`j' fitm`z'_`j'
	}
}

*Categorical outcomes: alcohol and physical activity
clear
use "F:\Smums\Final analysis\bmjfinalsmumsdata",clear

*Produce frequency table of alcohol consumption by intervention group at baseline and 12 months.
tab  m0_alcohol_3_values treatment if m12_alcohol_3_values~=.
tab  m12_alcohol_3_values treatment if m0_alcohol_3_values~=.
*Conduct ordinal logistic regression of alcohol consumption by intervention group.
ologit m12_alcohol_3_values  treatment if  m0_alcohol_3_values~=.,or
*Conduct ordinal logistic regression of alcohol consumption by intervention group, adjusting for site, recruitment pathway and ethnicity.
ologit m12_alcohol_3_values treatment i.site i.recruitmentpathway_subgroups i.bl_ethnorg if m0_alcohol_3_values~=.,or

*Produce frequency table of activity level by intervention group at baseline and 12 months.
tab  m0_activitylevel treatment if m12_activitylevel~=.
tab  m12_activitylevel treatment if m0_activitylevel~=.
*Conduct ordinal logistic regression of activity level by intervention group.
ologit m12_activitylevel  treatment if  m0_activitylevel~=.,or
*Conduct ordinal logistic regression of activity level by intervention group, adjusting for site, recruitment pathway and ethnicity.
ologit m12_activitylevel treatment i.site i.recruitmentpathway_subgroups i.bl_ethnorg if m0_activitylevel~=.,or

*Produce frequency table of alcohol consumption by intervention group at baseline and 6 months.
tab  m0_alcohol_3_values treatment if m6_alcohol_3_values~=.
tab  m6_alcohol_3_values treatment if m0_alcohol_3_values~=.
*Conduct ordinal logistic regression of alcohol consumption by intervention group.
ologit m6_alcohol_3_values  treatment if  m0_alcohol_3_values~=.,or
*Conduct ordinal logistic regression of alcohol consumption by intervention group, adjusting for site, recruitment pathway and ethnicity.
ologit m6_alcohol_3_values treatment i.site i.recruitmentpathway_subgroups i.bl_ethnorg if m0_alcohol_3_values~=.,or

*Produce frequency table of activity level by intervention group at baseline and 6 months.
tab  m0_activitylevel treatment if m6_activitylevel~=.
tab  m6_activitylevel treatment if m0_activitylevel~=.
*Conduct ordinal logistic regression of activity level by intervention group.
ologit m6_activitylevel  treatment if  m0_activitylevel~=.,or
*Conduct ordinal logistic regression of activity level by intervention group, adjusting for site, recruitment pathway and ethnicity.
ologit m6_activitylevel treatment i.site i.recruitmentpathway_subgroups i.bl_ethnorg if m0_activitylevel~=.,or


**Table 4:Comparisons of weight (kg) between intervention and active control groups at 12 months by prespecified subgroups.
clear
*Conduct analysis in subgroups defined by site (site), recruitment pathway (recruitmentpathway_subgroups), ethnicity (bl_ethnorg), deprivation (imd_quintile), employment (bl_empl_recode), education (bl_education), income (bl_income_30andunder), baseline BMI (bl_bmi_categories), parity (bl_parity), weeks postpartum (blpostpart)
foreach j in site recruitmentpathway_subgroups bl_ethnorg  imd_quintile bl_empl_recode bl_education bl_income_30andunder bl_bmi_categories  bl_parity blpostpart {
use "F:\Smums\Final analysis\bmjfinalsmumsdata",clear
drop if `j'==.		
*Recode education variable to 5 categories.
replace bl_education=2 if bl_education==1
*Conduct ANCOVA with interaction between treatment and subgroup variable to get P for interaction.
regress  m12_wght_kg i.treatment bl_wght_kg i.treatment##i.`j'
testparm i.treatment#i.`j'
*Conduct ANCOVA including site, recruitment pathway and ethnicity and interaction between treatment and subgroup variable to get adjusted P for interaction.
regress  m12_wght_kg i.treatment i.site i.recruitmentpathway_subgroups i.bl_ethnorg bl_wght_kg i.treatment##i.`j'
testparm i.treatment#i.`j'

decode `j',gen(lab`j')
replace `j'=9999 if `j'==.
levelsof `j',local(levels)	
	foreach i of local levels{
	*Calculate summary statistics within each subgroup.
	summarize  bl_wght_kg if treatment==0 & m12_wght_kg~=. & bl_wght_kg~=. & `j'==`i'
	summarize  bl_wght_kg if treatment==1 & m12_wght_kg~=. & bl_wght_kg~=. & `j'==`i'
	summarize  m12_wght_kg if treatment==0 & m12_wght_kg~=. & bl_wght_kg~=. & `j'==`i'
	summarize  m12_wght_kg if treatment==1 & m12_wght_kg~=. & bl_wght_kg~=. & `j'==`i'
	*Conduct ANCOVA to calculate difference in mean by intervention group (and 95% CI and P value) adjusting for baseline within each subgroup.
	regress  m12_wght_kg i.treatment bl_wght_kg if `j'==`i'
	*Conduct ANCOVA to calculate difference in mean by intervention group (and 95% CI and P value) adjusting for baseline and site, recruitment pathway and ethnicity, within each subgroup. 
	regress  m12_wght_kg i.treatment i.site i.recruitmentpathway_subgroups i.bl_ethnorg bl_wght_kg if `j'==`i'
	}
}


**Supplementary Table 2: Infant feeding outcomes based on portions of different food items given to infants weekly comparing the intervention and active control groups at 12 months
clear
use "F:\Smums\Final analysis\bmjfinalsmumsdata",clear
	*Conduct analysis separately for each food listed in Supplementary Table 2.
	foreach j in bcereals_portions brice_portions bbread_portions bpot_portions bchips_portions bbutter_portions bmeat_portions bprocmeat_portions bpoultry_portions bfish_portions beggs_portions bbeans_portions btofu_portions bnuts_portions bfruits_portions bveg_portions bdairy_portions bpud_portions bbiscuits_portions bcrisps_portions bformula_portions {
	*Calculate summary statistics by intervention group for each food at 12 months. 
	summarize  m0_`j' if treatment==0 & m12_`j'~=. & m0_`j'~=. 
	summarize  m0_`j' if treatment==1 & m12_`j'~=. & m0_`j'~=. 
	summarize  m12_`j' if treatment==0 & m12_`j'~=. & m0_`j'~=. 
	summarize  m12_`j' if treatment==1 & m12_`j'~=. & m0_`j'~=. 
	*Conduct ANCOVA to calculate difference in mean by intervention group (and 95% CI and P value) adjusting for baseline for each food at 12 months.
	regress  m12_`j' i.treatment m0_`j'
	*Conduct ANCOVA to calculate difference in mean by intervention group (and 95% CI and P value) adjusting for baseline and site, recruitment pathway and ethnicity, for each food at 12 months. 	
	regress  m12_`j' i.treatment i.site i.recruitmentpathway_subgroups i.bl_ethnorg m0_`j'
	}
	
**Supplementary Table 3: Infant feeding outcomes based on portions of different food items given to infants weekly comparing the intervention and active control groups at 6 months
clear
use "F:\Smums\Final analysis\bmjfinalsmumsdata",clear
	*Conduct analysis separately for each food listed in Supplementary Table 3.
	foreach j in bcereals_portions brice_portions bbread_portions bpot_portions bchips_portions bbutter_portions bmeat_portions bprocmeat_portions bpoultry_portions bfish_portions beggs_portions bbeans_portions btofu_portions bnuts_portions bfruits_portions bveg_portions bdairy_portions bpud_portions bbiscuits_portions bcrisps_portions bformula_portions {
	*Calculate summary statistics by intervention group for each food at 6 months. 
	summarize  m0_`j' if treatment==0 & m6_`j'~=. & m0_`j'~=. 
	summarize  m0_`j' if treatment==1 & m6_`j'~=. & m0_`j'~=. 
	summarize  m6_`j' if treatment==0 & m6_`j'~=. & m0_`j'~=. 
	summarize  m6_`j' if treatment==1 & m6_`j'~=. & m0_`j'~=. 
	*Conduct ANCOVA to calculate difference in mean by intervention group (and 95% CI and P value) adjusting for baseline for each food at 6 months.
	regress  m6_`j' i.treatment m0_`j'
	*Conduct ANCOVA to calculate difference in mean by intervention group (and 95% CI and P value) adjusting for baseline and site, recruitment pathway and ethnicity, for each food at 6 months. 	
	regress  m6_`j' i.treatment i.site i.recruitmentpathway_subgroups i.bl_ethnorg m0_`j'
	}

**Supplementary Table 4: Comparisons of FFB scores between intervention and active control groups at 12 months by prespecified subgroups.
clear
*Conduct analysis in subgroups defined by site (site), recruitment pathway (recruitmentpathway_subgroups), ethnicity (bl_ethnorg), deprivation (imd_quintile), employment (bl_empl_recode), education (bl_education), income (bl_income_30andunder), baseline BMI (bl_bmi_categories), parity (bl_parity), weeks postpartum (blpostpart)
foreach j in site recruitmentpathway_subgroups bl_ethnorg  imd_quintile bl_empl_recode bl_education bl_income_30andunder bl_bmi_categories  bl_parity blpostpart {
use "F:\Smums\Final analysis\bmjfinalsmumsdata",clear
drop if `j'==.	
*Recode education variable to 5 categories.	
replace bl_education=2 if bl_education==1
*Conduct ANCOVA with interaction between treatment and subgroup variable to get P for interaction.
regress  m12_ffbscore i.treatment m0_ffbscore i.treatment##i.`j'
testparm i.treatment#i.`j'
*Conduct ANCOVA including site, recruitment pathway and ethnicity and interaction between treatment and subgroup variable to get adjusted P for interaction.
regress  m12_ffbscore i.treatment i.site i.recruitmentpathway_subgroups i.bl_ethnorg m0_ffbscore i.treatment##i.`j'
testparm i.treatment#i.`j'

decode `j',gen(lab`j')
replace `j'=9999 if `j'==.
levelsof `j',local(levels)	
	foreach i of local levels{
	*Calculate summary statistics within each subgroup.
	summarize  m0_ffbscore if treatment==0 & m12_ffbscore~=. & m0_ffbscore~=. & `j'==`i'
	summarize  m0_ffbscore if treatment==1 & m12_ffbscore~=. & m0_ffbscore~=. & `j'==`i'
	summarize  m12_ffbscore if treatment==0 & m12_ffbscore~=. & m0_ffbscore~=. & `j'==`i'
	summarize  m12_ffbscore if treatment==1 & m12_ffbscore~=. & m0_ffbscore~=. & `j'==`i'
	*Conduct ANCOVA to calculate difference in mean by intervention group (and 95% CI and P value) adjusting for baseline within each subgroup.
	regress  m12_ffbscore i.treatment m0_ffbscore if `j'==`i'
	*Conduct ANCOVA to calculate difference in mean by intervention group (and 95% CI and P value) adjusting for baseline and site, recruitment pathway and ethnicity, within each subgroup. 
	regress  m12_ffbscore i.treatment i.site i.recruitmentpathway_subgroups i.bl_ethnorg m0_ffbscore if `j'==`i'
	}
}

**Supplementary Table 5: Comparisons of IPAQ-SF Total MET minutes/week between intervention and active control groups at 12 months by prespecified subgroups.
clear
*Conduct analysis in subgroups defined by site (site), recruitment pathway (recruitmentpathway_subgroups), ethnicity (bl_ethnorg), deprivation (imd_quintile), employment (bl_empl_recode), education (bl_education), income (bl_income_30andunder), baseline BMI (bl_bmi_categories), parity (bl_parity), weeks postpartum (blpostpart)
foreach j in site recruitmentpathway_subgroups bl_ethnorg  imd_quintile bl_empl_recode bl_education bl_income_30andunder bl_bmi_categories  bl_parity blpostpart {
use "F:\Smums\Final analysis\bmjfinalsmumsdata",clear
drop if `j'==.
*Recode education variable to 5 categories.	
replace bl_education=2 if bl_education==1
*Conduct ANCOVA with interaction between treatment and subgroup variable to get P for interaction.
regress  m12_totalpa_metmins i.treatment m0_totalpa_metmins i.treatment##i.`j'
testparm i.treatment#i.`j'
*Conduct ANCOVA including site, recruitment pathway and ethnicity and interaction between treatment and subgroup variable to get adjusted P for interaction.
regress  m12_totalpa_metmins i.treatment i.site i.recruitmentpathway_subgroups i.bl_ethnorg m0_totalpa_metmins i.treatment##i.`j'
testparm i.treatment#i.`j'

decode `j',gen(lab`j')
replace `j'=9999 if `j'==.
levelsof `j',local(levels)	
	foreach i of local levels{
	*Calculate summary statistics within each subgroup.
	summarize  m0_totalpa_metmins if treatment==0 & m12_totalpa_metmins~=. & m0_totalpa_metmins~=. & `j'==`i'
	summarize  m0_totalpa_metmins if treatment==1 & m12_totalpa_metmins~=. & m0_totalpa_metmins~=. & `j'==`i'
	summarize  m12_totalpa_metmins if treatment==0 & m12_totalpa_metmins~=. & m0_totalpa_metmins~=. & `j'==`i'
	summarize  m12_totalpa_metmins if treatment==1 & m12_totalpa_metmins~=. & m0_totalpa_metmins~=. & `j'==`i'
	*Conduct ANCOVA to calculate difference in mean by intervention group (and 95% CI and P value) adjusting for baseline within each subgroup.
	regress  m12_totalpa_metmins i.treatment m0_totalpa_metmins if `j'==`i'
	*Conduct ANCOVA to calculate difference in mean by intervention group (and 95% CI and P value) adjusting for baseline and site, recruitment pathway and ethnicity, within each subgroup. 
	regress  m12_totalpa_metmins i.treatment i.site i.recruitmentpathway_subgroups i.bl_ethnorg m0_totalpa_metmins if `j'==`i'
	}
}

**Supplementary Table 6:Views of participants on trial processes based on questionnaire responses at 12 months and feedback from qualitative interviews at 6 and 12 months
**Information
*Overall
cii prop 658 643
cii prop 658 10
cii prop 658 5
*Intervention
cii prop 311 304
cii prop 311 4
cii prop 311 3
*Control
cii prop 347 339
cii prop 347 6
cii prop 347 2
**Researcher
*Overall
cii prop 622 587
cii prop 622 24
cii prop 622 11
*Intervention
cii prop 294 279
cii prop 294 11
cii prop 294 4
*Control
cii prop 328 308
cii prop 328 13
cii prop 328 7
**Location
*Overall
cii prop 655 634
cii prop 655 19
cii prop 655 2
*Intervention
cii prop 309 294
cii prop 309 14
cii prop 309 1
*Control
cii prop 346 340
cii prop 346 5
cii prop 346 1
**Length of visit
*Overall
cii prop 654 641
cii prop 654 13
cii prop 654 0
*Intervention
cii prop 309 299
cii prop 309 10
cii prop 309 0
*Control
cii prop 345 342
cii prop 345 3
cii prop 345 0
**Weight taken 
*Overall
cii prop 655 634
cii prop 655 17
cii prop 655 4
*Intervention
cii prop 309 297
cii prop 309 10
cii prop 309 2
*Control
cii prop 346 337
cii prop 346 7
cii prop 346 2
**Height taken 
*Overall
cii prop 653 627
cii prop 653 23
cii prop 653 3
*Intervention
cii prop 308 296
cii prop 308 11
cii prop 308 1
*Control
cii prop 345 331
cii prop 345 12
cii prop 345 2
**Waist taken 
*Overall
cii prop 655 633
cii prop 655 18
cii prop 655 4
*Intervention
cii prop 309 297
cii prop 309 9
cii prop 309 3
*Control
cii prop 346 336
cii prop 346 9
cii prop 346 1
**Completing questions
*Overall
cii prop 653 573
cii prop 653 67
cii prop 653 13
*Intervention
cii prop 307 263
cii prop 307 38
cii prop 307 6
*Control
cii prop 346 310
cii prop 346 29
cii prop 346 7


**Supplementary Table 7: Questionnaire distribution and response rates by completion method, group, site, ethnicity and IMD quintile, at baseline, 6 and 12 months
*Baseline
tab bl_questreceived
tab bl_questmethod
tab bl_questmethod bl_questreceived,row
tab bl_questassist
tab treatment bl_questreceived,row 
tab site  bl_questreceived,row 
tab bl_ethnorg bl_questreceived,row 
tab imd_quintile bl_questreceived,row 
*6 months
tab m6_questreceived
tab m6_questmethod
tab m6_questmethod m6_questreceived,row
tab m6_questassist
tab treatment m6_questreceived,row 
tab site m6_questreceived,row 
tab bl_ethnorg m6_questreceived,row 
tab imd_quintile m6_questreceived,row 
*12 months
tab m12_questreceived
tab m12_questmethod
tab m12_questmethod m12_questreceived,row
tab m12_questassist
tab treatment m12_questreceived,row 
tab site m12_questreceived,row 
tab bl_ethnorg m12_questreceived,row 
tab imd_quintile m12_questreceived,row 


**Supplementary Figure 1.  Forest plot of adjusted mean difference in weight (and 95% confidence intervals) between intervention and active control group at 12 months by site.
clear
use "F:\Smums\Final analysis\bmjfinalsmumsdata",clear

postfile results str10 group bnusual bmeanusual bsdusual bnint bmeanint bsdint  anusual ameanusual asdusual anint ameanint asdint adjdiffmean adjdiffmeanlci adjdiffmeanuci adjdiffmeanp using "forestplot",replace

drop if site==.		
summarize  bl_wght_kg if treatment==0 & m12_wght_kg~=. & bl_wght_kg~=. 
local bnusual=r(N)
local bmeanusual =r(mean)
local bsdusual=r(sd)
summarize  bl_wght_kg if treatment==1 & m12_wght_kg~=. & bl_wght_kg~=. 
local bnint=r(N)
local bmeanint=r(mean)
local bsdint=r(sd)
summarize  m12_wght_kg if treatment==0 & m12_wght_kg~=. & bl_wght_kg~=. 
local anusual=r(N)
local ameanusual =r(mean)
local asdusual=r(sd)
summarize  m12_wght_kg if treatment==1 & m12_wght_kg~=. & bl_wght_kg~=.
local anint=r(N)
local ameanint=r(mean)
local asdint=r(sd)
regress  m12_wght_kg i.treatment i.site i.recruitmentpathway_subgroups i.bl_ethnorg bl_wght_kg
matrix f=r(table)
local adjdiffmean=f[1,2]
local adjdiffmeanlci= f[5,2]
local adjdiffmeanuci= f[6,2]
local adjdiffmeanp=f[4,2]
matrix drop f	
post results  ("overall") (`bnusual') (`bmeanusual') (`bsdusual') (`bnint') (`bmeanint') (`bsdint') (`anusual') (`ameanusual') (`asdusual') (`anint') (`ameanint') (`asdint') (`adjdiffmean') (`adjdiffmeanlci') (`adjdiffmeanuci') (`adjdiffmeanp')  

decode site,gen(labsite)
replace site=9999 if site==.
levelsof site,local(levels)	
	foreach i of local levels{
	summarize  bl_wght_kg if treatment==0 & m12_wght_kg~=. & bl_wght_kg~=. & site==`i'
	local bnusual=r(N)
	local bmeanusual =r(mean)
	local bsdusual=r(sd)
	summarize  bl_wght_kg if treatment==1 & m12_wght_kg~=. & bl_wght_kg~=. & site==`i'
	local bnint=r(N)
	local bmeanint=r(mean)
	local bsdint=r(sd)
	summarize  m12_wght_kg if treatment==0 & m12_wght_kg~=. & bl_wght_kg~=. & site==`i'
	local anusual=r(N)
	local ameanusual =r(mean)
	local asdusual=r(sd)
	summarize  m12_wght_kg if treatment==1 & m12_wght_kg~=. & bl_wght_kg~=. & site==`i'
	local anint=r(N)
	local ameanint=r(mean)
	local asdint=r(sd)
	regress  m12_wght_kg i.treatment i.site i.recruitmentpathway_subgroups i.bl_ethnorg bl_wght_kg if site==`i'
	matrix f=r(table)
	local adjdiffmean=f[1,2]
	local adjdiffmeanlci= f[5,2]
	local adjdiffmeanuci= f[6,2]
	local adjdiffmeanp=f[4,2]
	matrix drop f	

	post results  ("site`i'") (`bnusual') (`bmeanusual') (`bsdusual') (`bnint') (`bmeanint') (`bsdint') (`anusual') (`ameanusual') (`asdusual') (`anint') (`ameanint') (`asdint') (`adjdiffmean') (`adjdiffmeanlci') (`adjdiffmeanuci') (`adjdiffmeanp')  
	}
postclose results
clear
use "forestplot"

gen newgroup=1 if group=="site1"
replace newgroup=2 if group=="site2"
replace newgroup=3 if group=="site3"
replace newgroup=4 if group=="site4"
replace newgroup=5 if group=="site5"
replace newgroup=6.5 if group=="overall"

gen name="Belfast" if group=="site1"
replace name="Bradford"  if group=="site2"
replace name="London"  if group=="site3"
replace name="Scotland"  if group=="site4"
replace name="Cardiff"  if group=="site5"
replace name="Overall estimate" if group=="overall"

gen labx=-8

graph twoway (scatter  newgroup adjdiffmean if newgroup~=6.5, msymbol(square) mcolor(black) yscale(off reverse range(1 7)) ylabel(none) xline(0, lcolor(black) lpattern(dot)) xtitle("                               Adjusted mean difference (95% CI)") xscale(range(-8,4)) xlabel(-4 -2 0 2 4, nogrid) plotregion(c(white style(none))) graphregion(c(white) style(none)) legend(off)) ///
			(rcap adjdiffmeanlci adjdiffmeanuci newgroup if newgroup~=6.5, lcolor(black) horizontal msize(0.5)) ///
			(scatter  newgroup adjdiffmean if newgroup==6.5, msymbol(diamond) mcolor(black)) ///
			(rcap adjdiffmeanlci adjdiffmeanuci newgroup if newgroup==6.5, lcolor(black) horizontal msize(0.5)) ////
			 (scatter newgroup labx,mlabel(name) mlabcolor(black) msymbol(none) mlabsize(medium))

log close
