## Supplementary information for "Effectiveness of an automated text message intervention for weight management in postpartum women with overweight or obesity (Supporting MumS (SMS)): a UK wide, multicentre, two arm, parallel group, randomised controlledti trial"

**Acceptability of trial methods**

***Methods***

Assessment of the acceptability of the trial methods with the diverse sample of women from across the UK recruited to the SMS trial was conducted as part of the trial process evaluation (see PEAP in Supplementary information), to inform future research. Acceptability of the trials methods for participants was assessed using data recorded in the CRF or trial records at each time point. These included location of data collection visits (home, university or other), visit timing and date, questionnaire completion method (paper or online), level of assistance for questionnaire completion (researcher-assisted or independent), and questionnaire return status. Instances of declined measurements were documented in the CRF notes.

In the 12 month questionnaires, participants rated trial methods using 5 point scales, as follows: 1) agreement (*strongly agree* to *strongly disagree)* with statements, such as ‘*at the start of the study, the information given about the study was clear and informative’*; 2) level of difficulty (*very easy* to *very difficult)* with location and length of data collection visits, having weight, height and waist circumference measured and completing questionnaires. Optional free text responses were captured to explain ratings.

***Analysis***

At each timepoint, trial methods were summarised using numbers and proportions (n (%)) according to data collection visits by location (home, university, other), time of day (pre/post 17:00) and day (weekday/weekend (computed from date of visit)) as well as questionnaire completion method (paper/online), response receipt, and researcher assistance. Questionnaire response rates were examined by site, IMD quintile, ethnicity, allocation, and completion method. Free text ‘other’ locations were categorised. Participants declining weight or waist measurements were classified as decliners at each timepoint and numbers reported.

Questionnaire ratings of trial methods acceptability were summarised as frequencies (n, %) on five-point scales. Due to skew, responses were collapsed into three categories combining agreeable and disagreeable responses (e.g. ‘strongly agree’ and ‘agree’ combined into ‘strongly agree/agree’). Frequencies (n ,%) with binomial 95% CIs were then calculated by total sample and allocated group, to indicate overall acceptability.

***Results***

Trial methods were highly acceptable across the sample (Supplementary Table 6). Most visits occurred in participants’ homes (baseline: 85%; 6 months: 88%; 12 months: 90%) with few held in university settings (2-4%) and other locations (8-11%) including workplaces and community venues (e.g. centres, libraries, cafés). Across four sites (Belfast, Bradford, Scotland, Cardiff), home visits predominated (88–100%). London differed, with fewer home visits (53–66%) and more conducted in community settings. Visits were scheduled between 08:30 and 19:30; most started before 17:00 (>96%) and were conducted on weekdays (>92%). Questionnaire return rates were high (baseline: 97%; 6 months: 94%; 12 months: 95%) and consistent across participant characteristics, randomised groups and completion method (Supplementary Table 7). A small number declined weight (1 at 6 months: 1 at 12 months) or waist circumference measurements (2 at 6 months; 14 at 12 months), all at the Cardiff site.
