## Supplementary Tables 1 to 7 for "Effectiveness of an automated text message intervention for weight management in postpartum women with overweight or obesity (Supporting MumS (SMS)): a UK wide, multicentre, two arm, parallel group, randomised controlledti trial"

**Supplementary Table 1: A sample of intervention text messages as delivered over weeks 18 and 19 (days 120 to 133) of the intervention.**

| **Day of week** | **Day of intervention** | **Text message** |
| --- | --- | --- |
| Monday | 120 | Welcome to WEEK 18! Weight loss is much easier with support. If you have someone close to you, they can receive the same messages as you and support you on your journey. To do this, text 'SUPPORT' followed by the person's mobile number (ask their permission first). E.g. SUPPORT 07800000000. |
| Tuesday | 121 | Increase the fibre in your food to keep you feeling fuller for longer. Try some high fibre foods and introduce them gradually to your meals: high fibre cereal, wholemeal bread, and beans/lentils of all varieties. And don't forget to drink lots of water :) |
| Wednesday | 122 | How are you finding weekends? Are you able to stick to your diet and activity plans? Text Y15 for yes or N15 for no. |
| Friday | 124 | Friendly weight reminder text :) Text us, e.g. 11 st 5 or 73.3 kg. All texts you send are confidential :) |
| Saturday | 125 | Mum's tip: 'I made up every excuse under the sun when it came to exercise, so I signed up for a 6 week block of classes. I felt I had to go each week as I'd invested in myself. It worked - I felt good after it and I only missed one class'. Why not have a look at the fitness classes near you to get motivated? - <https://www.classfinder.org.uk/> |
| Sunday | 126 | Wondering how long until you see results?! You may enjoy some benefits of activity straight away e.g. happier mood, or even sleeping better. For changes in your heart and lung function, it can take up to 6 weeks. To notice changes in your clothes, it can take a little bit longer - but it is worth it, so keep going :) |
| Sunday | 126 | As this week comes to an end, focus on all the things that you do well! You have brought another little person into this world so give yourself the praise you deserve :) |
| Monday | 127 | Appreciate yourself in WEEK 19. Think of a time you achieved something you were proud of and how it made you feel. Keep this is mind for this journey :) |
| Tuesday | 128 | Smart snack ideas: apple slices spread with a thin layer of peanut butter (or just sprinkled with cinnamon), frozen grapes, some pineapple, or chopped carrots and peppers with a couple of spoonfuls of hummus (find a quick and easy recipe for hummus here - <https://www.bbcgoodfood.com/recipes/hummus> |
| Wednesday | 129 | Eating well doesn't have to break the bank. Check out these tips on how to eat well for less - <https://www.bda.uk.com/resource/food-facts-eat-well-spend-less.html> |
| Thursday | 130 | Squeezing in a quick workout (no matter how short) will boost your weight loss! Try skipping or using a hula hoop for 30 secs, rest for 30 secs and repeat as often as you can in 5 or 10 minutes. You could burn up to 50 calories in 5 minutes. If you don't have either, ask your friends or try a workout that doesn’t require equipment, like these aerobic videos - <https://www.youtube.com/watch?v=a44ayeoSfKM&list=PLnhASgDToTkutrA50HmDExSWITajnTuXG> |
| Friday | 131 | Well done on stepping on those scales for another week! Text us your weight e.g. 11 st 5 or 73.3 kg. |
| Saturday | 132 | Can't fight that craving, or just having a bad day in general? Remember you can text 'CRAVE' or 'BAD DAY' at anytime for instant support and advice. |
| Sunday | 133 | Tummy toning tip: Baby amused on the floor? Set the timer on your phone and try a 30 second plank beside them. Challenge yourself to a longer plank each day. |
| Sunday | 133 | The end of another week! Take time to reflect on how your goals went this week. Things didn't go as planned? Don't dwell on it, let that motivate you to do better next week! |

**Supplementary Table 2: Infant feeding outcomes based on portions of different food items given to infants weekly comparing the intervention and active control groups at 12 months**

|  | Intervention | | | Active Control | | | Diff in mean^b^  (95% CI) | *P* | Adjusted^c^ diff in mean (95% CI) | *P* |
| --- | --- | --- | --- | --- | --- | --- | --- | --- | --- | --- |
|  | N^a^ | Baseline | Endpoint | N^a^ | Baseline | Endpoint |  |  |  |  |
|  |  | Mean (sd) | Mean (sd) |  | Mean (sd) | Mean (sd) |  |  |  |  |
| Weekly portions of infant food items at 12 months | | | | | | | | | | |
| Breakfast cereals | 206 | 4.84 (3.12) | 4.91 (2.80) | 227 | 4.81 (2.98) | 5.14 (2.88) | -0.23 (-0.76 to 0.29) | 0.38 | -0.18 (-0.71 to 0.34) | 0.49 |
| Rice or pasta | 207 | 2.91 (2.43) | 3.46 (2.27) | 225 | 2.58 (2.19) | 3.22 (2.32) | 0.14 (-0.27 to 0.55) | 0.51 | 0.24 (-0.16 to 0.65) | 0.24 |
| Bread | 205 | 3.39 (2.88) | 4.08 (2.76) | 222 | 3.19 (2.75) | 4.14 (2.72) | -0.12 (-0.62 to 0.39) | 0.65 | -0.17 (-0.67 to 0.33) | 0.51 |
| Potatoes | 206 | 2.25 (1.89) | 2.60 (2.13) | 227 | 2.27 (1.84) | 2.52 (1.92) | 0.09 (-0.28 to 0.45) | 0.64 | 0.12 (-0.24 to 0.49) | 0.51 |
| Potato products | 205 | 1.07 (1.72) | 1.78 (1.77) | 224 | 1.32 (1.92) | 1.94 (1.98) | -0.06 (-0.40 to 0.27) | 0.71 | -0.02 (-0.35 to 0.31) | 0.90 |
| Butter / margarine | 205 | 2.40 (2.90) | 3.01 (2.63) | 226 | 2.27 (2.77) | 3.15 (2.74) | -0.19 (-0.66 to 0.28) | 0.43 | -0.23 (-0.69 to 0.24) | 0.34 |
| Red meat | 202 | 1.07 (1.31) | 1.54 (1.69) | 224 | 1.09 (1.61) | 1.38 (1.39) | 0.17 (-0.12 to 0.45) | 0.25 | 0.22 (-0.07 to 0.50) | 0.14 |
| Processed meat | 204 | 0.68 (1.42) | 1.02 (1.44) | 225 | 0.59 (1.37) | 1.04 (1.43) | -0.06 (-0.31 to 0.18) | 0.62 | -0.11 (-0.35 to 0.13) | 0.36 |
| Chicken / poultry | 205 | 1.73 (1.45) | 2.37 (1.65) | 224 | 2.08 (1.86) | 2.38 (1.67) | 0.11 (-0.19 to 0.41) | 0.48 | 0.14 (-0.16 to 0.43) | 0.37 |
| Fish | 202 | 1.15 (1.17) | 1.54 (1.50) | 225 | 1.27 (1.42) | 1.46 (1.29) | 0.12 (-0.14 to 0.37) | 0.38 | 0.11 (-0.14 to 0.36) | 0.39 |
| Eggs | 204 | 1.46 (1.78) | 1.99 (2.10) | 220 | 1.66 (2.08) | 1.72 (1.90) | **0.37 (0.02 to 0.71)** | **0.04** | **0.47 (0.14 to 0.81)** | **0.01** |
| Beans / lentils | 205 | 1.40 (1.63) | 1.90 (1.71) | 223 | 1.52 (1.69) | 1.62 (1.68) | **0.32 (0.02 to 0.63)** | **0.04** | **0.40 (0.09 to 0.71)** | **0.01** |
| Tofu/ Quorn | 204 | 0.29 (0.91) | 0.43 (1.26) | 224 | 0.36 (1.06) | 0.40 (1.19) | 0.07 (-0.14 to 0.28) | 0.52 | 0.10 (-0.11 to 0.31) | 0.35 |
| Nuts | 200 | 0.57 (1.40) | 0.93 (1.82) | 226 | 0.57 (1.39) | 0.62 (1.39) | **0.31 (0.02 to 0.61)** | **0.04** | **0.35 (0.06 to 0.64)** | **0.02** |
| Fruits | 204 | 7.46 (3.35) | 8.47 (2.94) | 224 | 7.63 (3.26) | 8.35 (3.09) | 0.17 (-0.36 to 0.71) | 0.53 | 0.04 (-0.50 to 0.57) | 0.89 |
| Vegetables | 204 | 7.22 (3.47) | 7.46 (3.35) | 226 | 7.35 (3.31) | 7.11 (3.37) | 0.42 (-0.13 to 0.97) | 0.13 | 0.33 (-0.22 to 0.88) | 0.24 |
| Cheese / yoghurt | 206 | 5.15 (3.54) | 6.13 (3.25) | 225 | 5.25 (3.34) | 6.06 (3.13) | 0.10 (-0.47 to 0.67) | 0.74 | 0.01 (-0.55 to 0.56) | 0.99 |
| Puddings / desserts | 205 | 1.45 (2.28) | 1.69 (2.12) | 222 | 1.57 (2.33) | 1.88 (2.37) | -0.15 (-0.55 to 0.26) | 0.47 | -0.14 (-0.55 to 0.27) | 0.50 |
| Biscuits / sweets | 204 | 1.56 (2.35) | 2.88 (2.89) | 224 | 1.53 (2.28) | 2.80 (2.73) | 0.07 (-0.42 to 0.56) | 0.79 | 0.12 (-0.37 to 0.62) | 0.62 |
| Crisps and corn snacks | 205 | 1.92 (2.36) | 2.44 (2.26) | 225 | 1.94 (2.39) | 2.66 (2.33) | -0.22 (-0.63 to 0.19) | 0.29 | -0.16 (-0.57 to 0.25) | 0.45 |
| Follow-on formula | 203 | 1.74 (3.75) | 0.73 (2.29) | 226 | 2.19 (4.00) | 0.78 (2.45) | 0.02 (-0.42 to 0.45) | 0.94 | 0.10 (-0.33 to 0.54) | 0.64 |

^a^ At baseline, 258 participants (28.9%) (intervention n= 132 (29.7%); active control n= 126 (28.2%) had not yet introduced solid infant foods and were therefore excluded from the analysis at 12 months.

^b^ Using ANCOVA adjusting for baseline values.

^c^ Same as ^b^ but additionally adjusting for site, recruitment pathway (NHS or community) and ethnicity.

**Supplementary Table 3: Infant feeding outcomes based on portions of different food items given to infants weekly comparing the intervention and active control groups at 6 months**

|  | Intervention | | | Active Control | | | Diff in mean^b^  (95% CI) | *P* | Adjusted^c^ diff in mean (95% CI) | *P* |
| --- | --- | --- | --- | --- | --- | --- | --- | --- | --- | --- |
|  | N^a^ | Baseline | Endpoint | N^a^ | Baseline | Endpoint |  |  |  |  |
|  |  | Mean (sd) | Mean (sd) |  | Mean (sd) | Mean (sd) |  |  |  |  |
| Weekly portions of infant food items at 6 months | | | | | | | | | | |
| Breakfast cereals | 226 | 4.92 (3.10) | 4.99 (2.83) | 254 | 4.76 (3.01) | 5.12 (2.88) | -0.18 (-0.66 to 0.30) | 0.46 | -0.17 (-0.65 to 0.31) | 0.49 |
| Rice or pasta | 226 | 2.87 (2.41) | 3.31 (2.30) | 251 | 2.62 (2.26) | 3.10 (2.15) | 0.14 (-0.24 to 0.52) | 0.47 | 0.16 (-0.22 to 0.54) | 0.41 |
| Bread | 226 | 3.39 (2.88) | 4.07 (2.82) | 252 | 3.22 (2.78) | 4.01 (2.82) | 0.00 (-0.48 to 0.48) | 1.00 | -0.03 (-0.50 to 0.43) | 0.89 |
| Potatoes | 225 | 2.30 (1.93) | 2.47 (1.96) | 252 | 2.25 (1.84) | 2.25 (1.56) | 0.21 (-0.10 to 0.52) | 0.18 | 0.21 (-0.11 to 0.52) | 0.20 |
| Potato products | 221 | 1.07 (1.71) | 1.69 (2.06) | 248 | 1.35 (2.01) | 1.61 (1.85) | 0.20 (-0.12 to 0.52) | 0.22 | 0.21 (-0.11 to 0.53) | 0.20 |
| Butter / margarine | 224 | 2.40 (2.91) | 2.96 (2.91) | 253 | 2.31 (2.81) | 2.83 (2.65) | 0.10 (-0.37 to 0.56) | 0.69 | 0.06 (-0.40 to 0.53) | 0.79 |
| Red meat | 222 | 1.07 (1.37) | 1.17 (1.24) | 252 | 1.08 (1.59) | 1.17 (1.32) | -0.01 (-0.23 to 0.22) | 0.96 | -0.01 (-0.23 to 0.22) | 0.95 |
| Processed meat | 222 | 0.69 (1.45) | 0.89 (1.45) | 252 | 0.55 (1.26) | 0.80 (1.41) | 0.01 (-0.22 to 0.24) | 0.92 | 0.01 (-0.22 to 0.23) | 0.94 |
| Chicken / poultry | 223 | 1.80 (1.50) | 2.25 (1.65) | 250 | 2.07 (1.89) | 2.22 (1.50) | 0.12 (-0.14 to 0.39) | 0.37 | 0.14 (-0.12 to 0.41) | 0.29 |
| Fish | 222 | 1.17 (1.15) | 1.49 (1.46) | 252 | 1.26 (1.42) | 1.40 (1.20) | 0.12 (-0.11 to 0.35) | 0.32 | 0.10 (-0.13 to 0.33) | 0.38 |
| Eggs | 224 | 1.57 (1.88) | 1.85 (2.05) | 247 | 1.61 (1.98) | 1.63 (1.77) | 0.24 (-0.06 to 0.54) | 0.12 | 0.27 (-0.03 to 0.56) | 0.08 |
| Beans / lentils | 224 | 1.44 (1.65) | 1.83 (1.72) | 250 | 1.52 (1.63) | 1.75 (1.61) | 0.11 (-0.18 to 0.39) | 0.46 | 0.13 (-0.15 to 0.42) | 0.37 |
| Tofu/ Quorn | 224 | 0.32 (0.99) | 0.37 (1.07) | 249 | 0.38 (1.21) | 0.36 (0.87) | 0.03 (-0.13 to 0.20) | 0.72 | 0.05 (-0.11 to 0.22) | 0.53 |
| Nuts | 222 | 0.67 (1.56) | 0.86 (1.79) | 252 | 0.50 (1.31) | 0.60 (1.42) | 0.20 (-0.07 to 0.47) | 0.15 | 0.22 (-0.06 to 0.49) | 0.12 |
| Fruits | 223 | 7.26 (3.42) | 8.41 (2.96) | 251 | 7.50 (3.35) | 8.02 (3.16) | 0.49 (0.00 to 0.98) | 0.05 | 0.42 (-0.06 to 0.91) | 0.09 |
| Vegetables | 222 | 7.11 (3.46) | 7.65 (3.28) | 251 | 7.16 (3.43) | 7.32 (3.17) | 0.35 (-0.15 to 0.84) | 0.17 | 0.21 (-0.28 to 0.71) | 0.40 |
| Cheese / yoghurt | 226 | 5.02 (3.51) | 6.16 (3.41) | 251 | 5.35 (3.44) | 6.00 (3.25) | 0.30 (-0.24 to 0.84) | 0.27 | 0.25 (-0.29 to 0.78) | 0.36 |
| Puddings / desserts | 225 | 1.50 (2.29) | 1.82 (2.52) | 249 | 1.52 (2.31) | 1.90 (2.34) | -0.07 (-0.46 to 0.32) | 0.73 | -0.07 (-0.47 to, 0.32) | 0.71 |
| Biscuits / sweets | 223 | 1.69 (2.43) | 2.33 (2.68) | 251 | 1.56 (2.31) | 2.37 (2.59) | -0.10 (-0.52 to 0.32) | 0.64 | -0.07 (-0.49 to 0.36) | 0.76 |
| Crisps and corn snacks | 224 | 2.13 (2.45) | 2.39 (2.39) | 251 | 1.98 (2.42) | 2.33 (2.25) | 0.00 (-0.38 to 0.39) | 0.99 | 0.06 (-0.33 to 0.44) | 0.77 |
| Follow-on formula | 222 | 1.91 (3.84) | 1.99 (3.79) | 249 | 1.92 (3.75) | 1.44 (3.35) | 0.55 (-0.07 to 1.16) | 0.08 | **0.63 (0.02 to 1.24)** | **0.04** |

^a^ At baseline, 258 participants (28.9%) (intervention n= 132 (29.7%); active control n= 126 (28.2%) had not yet introduced solid infant foods and were therefore excluded from the analysis at 6 months.

^b^ Using ANCOVA adjusting for baseline values.

^c^ Same as ^b^ but additionally adjusting for site, recruitment pathway (NHS or community) and ethnicity.

**Supplementary Table 4: Comparisons of FFB scores between intervention and active control groups at 12 months by prespecified subgroups.**

| Subgroups | Intervention | | | | | Active control | | | | Diff in mean^a^  (95% CI) | *P* interaction | Adjusted^b^ diff in  mean (95% CI) | *P* interaction |
| --- | --- | --- | --- | --- | --- | --- | --- | --- | --- | --- | --- | --- | --- |
|  | N | Baseline |  | Endpoint |  | N | Baseline |  | Endpoint |  |  |  |  |
|  |  | Mean (SD) | | Mean (SD) | |  | Mean (SD) | | Mean (SD) |  |  |  |  |
| Site |  |  | |  | |  |  | |  |  | 0.92 |  | 0.89 |
| Belfast | 68 | 3.0 (0.4) | | 3.1 (0.4) | | 85 | 3.1 (0.4) | | 3.1 (0.4) | 0.1 (-0.0 to 0.2) |  | 0.1 (0.0 to 0.2) |  |
| Bradford | 61 | 3.0 (0.4) | | 3.2 (0.5) | | 67 | 3.1 (0.4) | | 3.2 (0.4) | 0.0 (-0.1 to 0.2) |  | 0.0 (-0.1 to 0.1) |  |
| London | 60 | 3.1 (0.4) | | 3.3 (0.5) | | 73 | 3.1 (0.4) | | 3.1 (0.4) | 0.1 (0.0 to 0.2) |  | 0.1 (0.0 to 0.3) |  |
| Scotland | 63 | 2.9 (0.3) | | 3.2 (0.4) | | 59 | 3.0 (0.4) | | 3.2 (0.4) | 0.1 (-0.0 to 0.2) |  | 0.1 (-0.0 to 0.2) |  |
| Cardiff | 51 | 3.2 (0.4) | | 3.2 (0.4) | | 51 | 3.1 (0.5) | | 3.1 (0.5) | 0.1 (-0.0 to 0.2) |  | 0.1 (-0.0 to 0.2) |  |
| Recruitment |  |  | |  | |  |  | |  |  | 0.50 |  | 0.57 |
| NHS | 17 | 3.1 (0.4) | | 3.2 (0.4) | | 16 | 3.0 (0.4) | | 3.2 (0.4) | 0.0 (-0.2 to 0.2) |  | 0.1 (-0.1 to 0.3) |  |
| Community | 286 | 3.0 (0.4) | | 3.2 (0.4) | | 319 | 3.1 (0.4) | | 3.1 (0.4) | 0.1 (0.0 to 0.1) |  | 0.1 (0.0 to 0.1) |  |
| Ethnicity |  |  | |  | |  |  | |  |  | 0.85 |  | 0.85 |
| White | 216 | 3.0 (0.4) | | 3.2 (0.4) | | 220 | 3.1 (0.4) | | 3.1 (0.4) | 0.1 (0.0 to 0.1) |  | 0.1 (0.0 to 0.1) |  |
| Mixed/Multiple/Other | 17 | 3.2 (0.4) | | 3.2 (0.5) | | 15 | 3.1 (0.4) | | 3.1 (0.4) | 0.1 (-0.1 to 0.3) |  | 0.1 (-0.1 to 0.3) |  |
| Asian/Asian British | 39 | 3.0 (0.3) | | 3.2 (0.4) | | 61 | 3.1 (0.4) | | 3.1 (0.4) | 0.1 (-0.1 to 0.2) |  | 0.1 (-0.1 to 0.2) |  |
| Black/Caribbean | 30 | 3.1 (0.4) | | 3.3 (0.5) | | 39 | 3.2 (0.4) | | 3.2 (0.4) | 0.2 (-0.0 to 0.3) |  | 0.1 (-0.1 to 0.3) |  |
| IMD |  |  | |  | |  |  | |  |  | 0.80 |  | 0.80 |
| 1^st^ quintile- most deprived | 92 | 3.0 (0.4) | | 3.2 (0.5) | | 106 | 3.1 (0.5) | | 3.2 (0.5) | 0.1 (-0.0 to 0.2) |  | 0.1 (-0.0 to 0.2) |  |
| 2^nd^ quintile | 64 | 3.0 (0.4) | | 3.2 (0.5) | | 66 | 3.1 (0.4) | | 3.1 (0.4) | 0.2 (0.0 to 0.3) |  | 0.2 (0.0 to 0.3) |  |
| 3^rd^ quintile | 47 | 3.0 (0.4) | | 3.1 (0.4) | | 47 | 3.0 (0.3) | | 3.0 (0.3) | 0.1 (-0.0 to 0.2) |  | 0.1 (-0.0 to 0.2) |  |
| 4^th^ quintile | 40 | 3.0 (0.4) | | 3.2 (0.4) | | 57 | 3.1 (0.4) | | 3.2 (0.4) | 0.1 (-0.1 to 0.2) |  | 0.1 (-0.0 to 0.2) |  |
| 5^th^ quintile- least deprived | 60 | 3.1 (0.3) | | 3.2 (0.4) | | 59 | 3.1 (0.4) | | 3.1 (0.4) | 0.1 (0.0 to 0.2) |  | 0.1 (0.0 to 0.3) |  |
| Employment |  |  | |  | |  |  | |  |  | 0.82 |  | 0.68 |
| Full time | 127 | 3.1 (0.4) | | 3.2 (0.4) | | 157 | 3.1 (0.4) | | 3.1 (0.4) | 0.1 (0.0 to 0.1) |  | 0.1 (0.0 to 0.1) |  |
| Part time | 101 | 3.0 (0.4) | | 3.2 (0.4) | | 101 | 3.1 (0.4) | | 3.1 (0.4) | 0.1 (0.0 to 0.2) |  | 0.1 (0.0 to 0.2) |  |
| Unemployed/student | 69 | 3.0 (0.4) | | 3.2 (0.5) | | 75 | 3.1 (0.4) | | 3.2 (0.5) | 0.1 (-0.0 to 0.2) |  | 0.1 (-0.0 to 0.2) |  |
| Education (finished) |  |  | |  | |  |  | |  |  | 0.07 |  | 0.09 |
| Up to secondary (GCSE) | 24 | 2.9 (0.4) | | 3.1 (0.5) | | 29 | 3.0 (0.4) | | 3.2 (0.4) | -0.1 (-0.3 to 0.1) |  | -0.1 (-0.4 to 0.1) |  |
| Secondary (A Level) | 27 | 2.8 (0.3) | | 3.0 (0.3) | | 31 | 3.1 (0.4) | | 3.2 (0.5) | 0.0 (-0.1 to 0.2) |  | 0.0 (-0.2 to 0.2) |  |
| Further education | 44 | 3.2 (0.4) | | 3.3 (0.5) | | 42 | 3.1 (0.5) | | 3.1 (0.5) | 0.2 (0.0 to 0.3) |  | 0.2 (0.0 to 0.3) |  |
| UG degree | 120 | 3.0 (0.4) | | 3.2 (0.4) | | 131 | 3.1 (0.4) | | 3.2 (0.4) | 0.1 (0.0 to 0.1) |  | 0.1 (0.0 to 0.1) |  |
| PG degree | 88 | 3.1 (0.4) | | 3.3 (0.4) | | 102 | 3.1 (0.4) | | 3.1 (0.4) | 0.2 (0.1 to 0.2) |  | 0.2 (0.1 to 0.2) |  |
| Income |  |  | |  | |  |  | |  |  | 0.53 |  | 0.63 |
| Less than £30,000 | 96 | 3.0 (0.4) | | 3.1 (0.5) | | 93 | 3.1 (0.4) | | 3.2 (0.5) | 0.1 (-0.0 to 0.2) |  | 0.1 (-0.0 to 0.2) |  |
| £30,001 or more | 186 | 3.1 (0.3) | | 3.2 (0.4) | | 219 | 3.1 (0.4) | | 3.1 (0.4) | 0.1 (0.0 to 0.2) |  | 0.1 (0.1 to 0.2) |  |
| Don't know | 21 | 3.0 (0.3) | | 3.3 (0.4) | | 23 | 3.1 (0.5) | | 3.3 (0.4) | 0.1 (-0.2 to 0.3) |  | 0.0 (-0.3 to 0.3) |  |
| BMI (at study entry) |  |  | |  | |  |  | |  |  | 0.58 |  | 0.61 |
| 25.0-29.9 | 120 | 3.0 (0.3) | | 3.2 (0.4) | | 143 | 3.1 (0.4) | | 3.1 (0.5) | 0.1 (0.0 to 0.2) |  | 0.1 (0.0 to 0.2) |  |
| 30.0 and over | 183 | 3.1 (0.4) | | 3.2 (0.5) | | 192 | 3.1 (0.4) | | 3.2 (0.4) | 0.1 (0.0 to 0.1) |  | 0.1 (0.0 to 0.1) |  |
| Parity |  |  | |  | |  |  | |  |  | 0.06 |  | 0.03 |
| 1 | 132 | 3.0 (0.4) | | 3.2 (0.4) | | 146 | 3.1 (0.4) | | 3.2 (0.4) | 0.0 (-0.0 to 0.1) |  | 0.0 (-0.0 to 0.1) |  |
| 2 | 115 | 3.0 (0.4) | | 3.2 (0.4) | | 112 | 3.1 (0.4) | | 3.1 (0.4) | 0.1 (0.0 to 0.2) |  | 0.1 (0.0 to 0.2) |  |
| 3+ | 56 | 3.0 (0.4) | | 3.3 (0.5) | | 77 | 3.1 (0.5) | | 3.1 (0.5) | 0.2 (0.1 to 0.3) |  | 0.2 (0.1 to 0.3) |  |
| Weeks postpartum (at study entry) |  |  | |  | |  |  | |  |  | 0.79 |  | 0.87 |
| ≤26 weeks (≤6 months) | 112 | 3.0 (0.4) | | 3.2 (0.4) | | 117 | 3.1 (0.4) | | 3.2 (0.5) | 0.1 (0.0 to 0.2) |  | 0.1 (0.0 to 0.2) |  |
| 27-52 weeks (7-12 months) | 84 | 3.1 (0.4) | | 3.2 (0.4) | | 100 | 3.1 (0.4) | | 3.1 (0.4) | 0.1 (-0.0 to 0.2) |  | 0.1 (-0.0 to 0.2) |  |
| ≥53 weeks (>12 months) | 107 | 3.0 (0.4) | | 3.2 (0.4) | | 118 | 3.1 (0.4) | | 3.2 (0.4) | 0.1 (0.0 to 0.2) |  | 0.1 (0.0 to 0.2) |  |

*^a^ Using ANCOVA (adjusting for baseline values).*

*^b^ Same as ^a^ but additionally adjusting for site, recruitment pathway (NHS or community) and ethnicity (using categories shown in Table 1).*

**Supplementary Table 5: Comparisons of IPAQ-SF Total MET minutes/week between intervention and active control groups at 12 months by prespecified subgroups.**

| Subgroups | Intervention | | | Active Control | | | Diff in mean^a^ (95% CI) | *P* interaction | Adjusted^b^ diff in mean (95% CI) | *P* interaction |
| --- | --- | --- | --- | --- | --- | --- | --- | --- | --- | --- |
|  | n | Baseline | Endpoint | N | Baseline | Endpoint |  |  |  |  |
|  |  | Mean (SD) | Mean (SD) |  | Mean (SD) | Mean (SD) |  |  |  |  |
| Site |  |  |  |  |  |  |  | 0.87 |  | 0.91 |
| Belfast | 67 | 1462 (1580) | 1932 (1958) | 85 | 1701 (1461) | 1688 (1618) | 340 (-203 to 882) |  | 363 (-190 to 915) |  |
| Bradford | 60 | 764 (921) | 1677 (1956) | 65 | 1047 (1198) | 1652 (1834) | 235 (-381 to 852) |  | 217 (-417 to 852) |  |
| London | 57 | 1750 (1469) | 2389 (2035) | 72 | 2010 (2021) | 1906 (2009) | 623 (-4 to 1249) |  | 493 (-161 to 1148) |  |
| Scotland | 62 | 1531 (1255) | 2077 (1765) | 59 | 1619 (1753) | 1688 (1686) | 421 (-170 to 1012) |  | 332 (-232 to 896) |  |
| Cardiff | 51 | 1691 (1337) | 2136 (1920) | 50 | 1613 (2306) | 1581 (1790) | 505 (-57 to 1067) |  | 485 (-97 to 1066) |  |
| Recruitment |  |  |  |  |  |  |  | 0.09 |  | 0.09 |
| NHS | 16 | 1750 (1088) | 2892 (2650) | 16 | 1560 (1353) | 1407 (1745) | 1411 (-206 to 3028) |  | 1378 (-603 to 3359) |  |
| Community | 281 | 1412 (1387) | 1985 (1874) | 315 | 1615 (1789) | 1728 (1785) | 361 (98 to 624) |  | 355 (90 to 620) |  |
| Ethnicity |  |  |  |  |  |  |  | 0.29 |  | 0.31 |
| White | 211 | 1531 (1381) | 2129 (1913) | 219 | 1686 (1772) | 1712 (1648) | 495 (193 to 798) |  | 473 (170 to 776) |  |
| Mixed/Multiple/Other | 17 | 1383 (1563) | 1614 (1652) | 15 | 2318 (2180) | 2766 (2540) | -467 (-1688 to 753) |  | -659 (-2054 to 737) |  |
| Asian/Asian British | 39 | 725 (747) | 1643 (1970) | 58 | 1069 (1415) | 1341 (1745) | 442 (-298 to 1182) |  | 402 (-374 to 1178) |  |
| Black/Caribbean | 29 | 1711 (1599) | 2179 (2119) | 39 | 1730 (1927) | 1859 (2092) | 329 (-630 to 1288) |  | -84 (-1002 to 834) |  |
| IMD |  |  |  |  |  |  |  | 0.78 |  | 0.77 |
| 1^st^ quintile- most deprived | 90 | 1446 (1609) | 2199 (2286) | 103 | 1697 (2344) | 1820 (1933) | 487 (-60 to 1035) |  | 535 (-25 to 1094) |  |
| 2^nd^ quintile | 61 | 1291 (1323) | 1832 (1746) | 65 | 1813 (1928) | 1891 (2334) | 300 (-319 to 919) |  | 184 (-454 to 822) |  |
| 3^rd^ quintile | 46 | 1253 (1290) | 2028 (2086) | 47 | 1594 (1115) | 1589 (1254) | 672 (41 to 1303) |  | 588 (-63 to 1239) |  |
| 4^th^ quintile | 40 | 1444 (1122) | 2141 (1613) | 57 | 1488 (1336) | 1565 (1388) | 588 (-6 to 1182) |  | 696 (65 to 1326) |  |
| 5^th^ quintile- least deprived | 60 | 1674 (1250) | 1922 (1592) | 59 | 1376 (1099) | 1568 (1503) | 164 (-334 to 662) |  | 194 (-346 to 734) |  |
| Employment |  |  |  |  |  |  |  | 0.58 |  | 0.64 |
| Full time | 124 | 1492 (1167) | 1926 (1808) | 157 | 1594 (1750) | 1490 (1542) | 497 (168 to 826) |  | 513 (179 to 847) |  |
| Part time | 99 | 1421 (1552) | 1971 (1769) | 99 | 1752 (2059) | 1793 (1803) | 301 (-166 to 768) |  | 286 (-183 to 755) |  |
| Unemployed/student | 69 | 1331 (1468) | 2198 (2207) | 73 | 1460 (1374) | 2128 (2144) | 152 (-509 to 813) |  | 118 (-547 to 783) |  |
| Education (finished) |  |  |  |  |  |  |  | 0.41 |  | 0.45 |
| Up to secondary (GCSE) | 23 | 922 (1749) | 2373 (2259) | 28 | 1747 (1642) | 2361 (2186) | 280 (-986 to 1546) |  | 76 (-1255 to 1407) |  |
| Secondary (A Level) | 26 | 1352 (1328) | 1469 (1387) | 31 | 1501 (1588) | 1923 (2144) | -359 (-1206 to 489) |  | -268 (-1230 to 694) |  |
| Further education | 43 | 1962 (1525) | 2722 (2651) | 41 | 1807 (2978) | 2001 (2262) | 636 (-284 to 1556) |  | 572 (-364 to 1508) |  |
| UG degree | 118 | 1356 (1197) | 2045 (1760) | 130 | 1629 (1451) | 1653 (1716) | 554 (165 to 944) |  | 597 (200 to 994) |  |
| PG degree | 87 | 1426 (1373) | 1757 (1689) | 101 | 1508 (1596) | 1427 (1307) | 363 (-35 to 760) |  | 333 (-81 to 748) |  |
| Income |  |  |  |  |  |  |  | 0.77 |  | 0.66 |
| Less than £30,000 | 92 | 1158 (1436) | 2179 (2258) | 91 | 1975 (2186) | 2068 (2279) | 504 (-121 to 1130) |  | 553 (-78 to 1183) |  |
| £30,001 or more | 185 | 1519 (1277) | 1933 (1706) | 218 | 1456 (1303) | 1532 (1409) | 369 (92 to 645) |  | 364 (83 to 646) |  |
| Don't know | 20 | 1853 (1749) | 2290 (2251) | 22 | 1661 (3215) | 2027 (2477) | 148 (-980 to 1276) |  | -266 (-1511 to 979) |  |
| BMI (at study entry) |  |  |  |  |  |  |  | 0.19 |  | 0.22 |
| 25.0-29.9 | 118 | 1560 (1461) | 2049 (1746) | 142 | 1596 (1635) | 1858 (1888) | 208 (-203 to 620) |  | 178 (-240 to 596) |  |
| 30.0 and over | 179 | 1344 (1308) | 2023 (2045) | 189 | 1624 (1868) | 1603 (1694) | 573 (234 to 913) |  | 536 (192 to 880) |  |
| Parity |  |  |  |  |  |  |  | 0.77 |  | 0.67 |
| 1 | 128 | 1434 (1403) | 1944 (1826) | 145 | 1695 (1899) | 1754 (1744) | 337 (-24 to 699) |  | 259 (-109 to 627) |  |
| 2 | 115 | 1435 (1319) | 1980 (1857) | 110 | 1505 (1633) | 1496 (1755) | 512 (61 to 963) |  | 478 (13 to 944) |  |
| 3+ | 54 | 1409 (1435) | 2358 (2288) | 76 | 1608 (1714) | 1945 (1877) | 526 (-128 to 1180) |  | 559 (-118 to 1237) |  |
| Weeks postpartum (at study entry) |  |  |  |  |  |  |  | 0.95 |  | 0.96 |
| ≤26 weeks (≤6 months) | 111 | 1325 (1403) | 1933 (1971) | 117 | 1537 (1881) | 1603 (1702) | 438 (10 to 866) |  | 418 (-22 to 857) |  |
| 27-52 weeks (7-12 months) | 82 | 1477 (1266) | 2051 (1985) | 99 | 1772 (1605) | 1854 (2032) | 360 (-187 to 907) |  | 409 (-146 to 964) |  |
| ≥53 weeks (>12 months) | 104 | 1505 (1427) | 2127 (1850) | 115 | 1551 (1793) | 1701 (1632) | 448 (35 to 861) |  | 303 (-110 to 715) |  |

*^a^ Using ANCOVA (adjusting for baseline values).*

*^b^ Same as ^a^ but additionally adjusting for site, recruitment pathway (NHS or community) and ethnicity (using categories shown in Table 1)*

**Supplementary Table 6: Views of participants on trial processes based on questionnaire responses at 12 months**

|  | **All** | | **Intervention** | | **Control** | |
| --- | --- | --- | --- | --- | --- | --- |
|  | **N** | **%  (95% CI)** | **N** | **%  (95% CI)** | **N** | **%  (95% CI)** |
| ***At the start of the study, the information I was given about the study was clear and informative*** | ***658*** |  | ***311*** |  | ***347*** |  |
| Strongly agree/agree | 643 | 97.7 (96.3 to 98.7) | 304 | 97.7 (95.4 to 99.1) | 339 | 97.7 (95.5 to 99.0) |
| Neither agree nor disagree | 10 | 1.5 (0.7 to 2.8) | 4 | 1.3 (0.4 to 3.3) | 6 | 1.7 (0.6 to 3.7) |
| Strongly disagree/disagree | 5 | 0.8 (0.2 to 1.8) | 3 | 1.0 (0.2 to 2.8) | 2 | 0.6 (0.1 to 2.1) |
| *Missing* | *1* |  | *1* |  | *0* |  |
| ***The amount of information and level of support provided by the researchers during the study was suited to my needs*** | ***622*** |  | ***294*** |  | ***328*** |  |
| Strongly agree/agree | 587 | 94.4 (92.3 to 96.0) | 279 | 94.9 (91.7 to 97.1) | 308 | 93.9 (90.7 to 96.2) |
| Neither agree nor disagree | 24 | 3.9 (2.5 to 5.7) | 11 | 3.7 (1.9 to 6.6) | 13 | 4.0 (2.1 to 6.7) |
| Strongly disagree/disagree | 11 | 1.8 (0.9 to 3.1) | 4 | 1.4 (0.4 to 3.4) | 7 | 2.1 (0.9 to 4.3) |
| *Missing* | *37* |  | *18* |  | *19* |  |
| ***Location of visits*** | ***655*** |  | ***309*** |  | ***346*** |  |
| Very easy/easy | 634 | 96.8 (95.1 to 98.0) | 294 | 95.1 (92.1 to 97.3) | 340 | 98.3 (96.3 to 99.4) |
| No strong opinion | 19 | 2.9 (1.8 to 4.5) | 14 | 4.5 (2.5 to 7.5) | 5 | 1.4 (0.5 to 3.3) |
| Very difficult/difficult | 2 | 0.3 (0.0 to 1.1) | 1 | 0.3 (0.0 to 1.8) | 1 | 0.3 (0.0 to 1.6) |
| *Missing* | *3* |  | *2* |  | *1* |  |
| ***Length of visits*** | ***654*** |  | ***309*** |  | ***345*** |  |
| Very easy/easy | 641 | 98.0 (96.6 to 98.9) | 299 | 96.8 (94.1 to 98.4) | 342 | 99.1 (97.5 to 99.8) |
| No strong opinion | 13 | 2.0 (1.1 to 3.4) | 10 | 3.2 (1.6 to 5.9) | 3 | 0.9 (0.2 to 2.5) |
| Very difficult/difficult | 0 | 0 (0.0 to 0.6) | 0 | 0 (0.0 to 1.2) | 0 | 0 (0.0 to 1.1) |
| *Missing* | *4* |  | *2* |  | *2* |  |
| ***Having weight taken*** | ***655*** |  | ***309*** |  | ***346*** |  |
| Very easy/easy | 634 | 97.0 (95.1 to 98.0) | 297 | 96.1 (93.3 to 98.0) | 337 | 97.4 (95.1 to 98.8) |
| No strong opinion | 17 | 2.6 (1.5 to 4.1) | 10 | 3.2 (1.6 to 5.9) | 7 | 2.0 (0.8 to 4.1) |
| Very difficult/difficult | 4 | 0.6 (0.2 to 1.6) | 2 | 0.6 (0.1 to 2.3) | 2 | 0.6 (0.1 to 2.1) |
| *Missing* | *3* |  | *2* |  | *1* |  |
| ***Having height measure taken*** | ***653*** |  | ***308*** |  | ***345*** |  |
| Very easy/easy | 627 | 96.0 (94.2 to 97.4) | 296 | 96.1 (93.3 to 98.0) | 331 | 95.9 (93.3 to 97.8) |
| No strong opinion | 23 | 3.5 (2.2 to 5.2) | 11 | 3.6 (1.8 to 6.3) | 12 | 3.5 (1.8 to 6.0) |
| Very difficult/difficult | 3 | 0.5 (0.1 to 1.3) | 1 | 0.3 (0.0 to 1.8) | 2 | 0.6 (0.1 to 2.1) |
| *Missing* | *3* |  | *2* |  | *1* |  |
| ***Having waist measure taken*** | ***655*** |  | ***309*** |  | ***346*** |  |
| Very easy/easy | 633 | 96.6 (95.0 to 97.9) | 297 | 96.1 (93.3 to 98.0) | 336 | 97.1 (94.7 to 98.6) |
| No strong opinion | 18 | 2.7 (1.6 to 4.3) | 9 | 2.9 (1.3 to 5.5) | 9 | 2.6 (1.2 to 4.9) |
| Very difficult/difficult | 4 | 0.6 (0.2 to 1.6) | 3 | 1.0 (0.2 to 2.8) | 1 | 0.3 (0.0 to 1.6) |
| *Missing* | *3* |  | *2* |  | *1* |  |
| ***Completing questionnaires*** | ***653*** |  | ***307*** |  | ***346*** |  |
| Very easy/easy | 573 | 87.7 (85.0 to 90.2) | 263 | 85.7 (81.2 to 89.4) | 310 | 89.6 (85.9 to 92.6) |
| No strong opinion | 67 | 10.3 (8.0 to 12.8) | 38 | 12.4 (8.9 to 16.6) | 29 | 8.4 (5.7 to 11.8) |
| Very difficult/difficult | 13 | 2.0 (1.1 to 3.4) | 6 | 2.0 (0.7 to 4.2) | 7 | 2.0 (0.8 to 4.1) |
| *Missing* | *5* |  | *4* |  | *1* |  |

**Supplementary Table 7: Questionnaire distribution and response rates** **by completion method, group, site, ethnicity and IMD quintile, at baseline, 6 and 12 months**

|  | Baseline | | 6 months | | 12 months | |
| --- | --- | --- | --- | --- | --- | --- |
|  | N (%) | | N (%) | | N (%) | |
| Questionnaires distributed | 892 | | 781 | | 692 | |
| Total responses received | 868 (97.3%) | | 733 (93.9%) | | 658 (95.1%) | |
|  | Requested | Responded | Requested | Responded | Requested | Responded |
| Paper completion | 396 (44.4%) | 387/396 (97.7%) | 220 (28.2%) | 205/220 (93.2%) | 159 (23.0%) | 149 (93.7%) |
| Qualtrics completion | 496 (55.6%) | 481/496 (97.0%) | 561 (71.8%) | 528/561 (94.1%) | 533 (77.0%) | 509 (95.5%) |
| Assistance from researcher | 54 (6.1%) | | 57 (7.3%) | | 29 (4.2%) | |
|  | Distributed | Responded | Distributed | Responded | Distributed | Responded |
| By Group |  |  |  |  |  |  |
| Intervention | 445 | 432 (97.1%) | 375 | 350 (93.3%) | 331 | 311 (94.0%) |
| Active Control | 447 | 436 (97.5%) | 406 | 383 (94.3%) | 361 | 347 (96.1%) |
| By Site |  |  |  |  |  |  |
| Belfast | 200 | 197 (98.5%) | 170 | 164 (96.5%) | 159 | 154 (96.9%) |
| Bradford | 194 | 193 (99.5%) | 167 | 152 (91.0%) | 144 | 131 (91.0%) |
| London | 189 | 181 (95.8%) | 171 | 159 (93.0%) | 147 | 138 (93.9%) |
| Scotland | 150 | 150 (100.0%) | 141 | 137 (97.2%) | 124 | 122 (98.4%) |
| Cardiff | 159 | 147 (92.5%) | 132 | 121 (91.7%) | 118 | 113 (95.8%) |
| By Ethnicity |  |  |  |  |  |  |
| White | 591 | 575 (97.3%) | 516 | 492 (95.3%) | 465 | 448 (96.3%) |
| Mixed/Multiple/Other | 49 | 49 (100.0%) | 45 | 43 (95.6%) | 35 | 34 (97.1%) |
| Asian/Asian British | 151 | 148 (98.0%) | 129 | 117 (90.7%) | 113 | 103 (91.2%) |
| Black/Black British/Caribbean/African | 100 | 95 (95.0%) | 90 | 80 (88.9%) | 78 | 72 (92.3%) |
| By IMD quintile |  |  |  |  |  |  |
| 1^st^ quintile (most deprived) | 299 | 291 (97.3%) | 257 | 235 (91.4%) | 222 | 203 (91.4%) |
| 2^nd^ quintile | 178 | 174 (97.8%) | 156 | 146 (93.6%) | 139 | 133 (95.7%) |
| 3^rd^ quintile | 124 | 121 (97.6%) | 111 | 107 (96.4%) | 103 | 98 (95.1%) |
| 4^th^ quintile | 135 | 128 (94.8%) | 119 | 114 (95.8%) | 104 | 102 (98.1%) |
| 5^th^ quintile (least deprived) | 154 | 152 (98.7%) | 137 | 130 (94.9%) | 124 | 122 (98.4%) |
